# Improving Medication Prescribing-Related Outcomes for Vulnerable Elderly In Transitions on High Risk Medications (IMPROVE-IT HRM): A Pilot Randomized Trial Protocol

**DOI:** 10.1101/2023.03.24.23287691

**Authors:** Anne Holbrook, Dan Perri, Mitch Levine, Sarah Jarmain, Lehana Thabane, Jean-Eric Tarride, Lisa Dolovich, Sylvia Hyland, Alan Forster, Carmine Nieuwstraten

## Abstract

**Rationale:** Transitions in, through, and out of hospital define the highest risk periods for patient safety. Hospitalized senior high-cost health care users taking high risk medications, are a large group of patients, usually highly complex with polypharmacy, and at high risk of serious adverse medication events. We will assess whether an expert Clinical Pharmacology Toxicology (CPT) medication management intervention during hospitalization with follow-up post-discharge and communication with circle of care, is feasible and can decrease drug therapy problems amongst this group.

**Design:** Pragmatic pilot randomized trial at SJHH with 1:1 patient-level concealed randomization with blinded outcome assessment and data analysis.

**Participants:** Adults 65 years of age and older, admitted to Internal Medicine services for more than 2 days, who are high-cost users defined as at least one other hospitalization in the prior year, taking 5 or more chronic medications including at least one high risk medication.

**Intervention:** CPT consult service identifies medication target(s), completes consult, including priorities for improving prescribing negotiated with the patient, starts the care plan, ensures a detailed discharge medication reconciliation and circle-of-care communication, and sees the patient at least twice after hospital discharge via integrated virtual visits to consolidate the care plan in the community. Control group receives usual care as provided by admitting services.

**Outcomes:** Include a) Feasibility Outcomes and b) Clinical Outcomes including the number of drug therapy problems improved, medication appropriateness and safety, the quality and coordination of transitions in care, quality of life, and health care utilization and costs by 3-month follow-up.

**Impact:** If results support feasibility of ramp-up and promising clinical outcomes, a follow-up definitive trial will be organized using a developing national platform and medication appropriateness network.

**RESEARCH QUESTION:** Our detailed research question is ‘In a randomized pilot trial, can an expert Clinical Pharmacology team coordinate and improve medication management during the very high-risk transition period from hospitalization through post-hospital discharge follow-up for senior high-cost users of healthcare taking high risk medications, meeting key feasibility outcomes while improving patient-important outcomes and health care costs sufficiently to warrant a large subsequent trial?’

## INTRODUCTION

Current systematic reviews of randomized trials to manage polypharmacy or to manage medications in hospital or in the transitions of care have been unable to demonstrate improvements in important clinical outcomes.^1–5^ This is largely because a) the interventions have been led by providers without the requisite combination of diagnostic, therapeutic and risk management expertise or authority to make requisite changes, b) the intervention was not sufficiently concentrated (too short, incomplete or misdirected), c) the medication focus was misplaced (i.e., not high-risk medications, which are the most associated with adverse clinical outcomes) or the outcomes were overly focused on poor quality surrogates for clinical outcomes.

Because of the huge potential burden of harm and the number of affected vulnerable older adults in our population (details below), the value of more randomized trial inquiry is <u>very</u> <u>high</u>. There is enough suggestion of benefit from trials on prescribing appropriateness combined with the potential for major medication safety improvements,^4, 6–9^ to support an expert intervention concentrated on the highest risk group as they transition through a very high-risk period targeting the highest risk medications. In addition, recent advances in digital health in Canada make it feasible for scarce clinical expertise to be concentrated on this high priority problem with robust communication systems that support patient consultation and follow-up to any part of country no matter how remote.

### Medication Safety is an International Health Priority

In 2017, the World Health Organization (WHO) declared Medication Safety to be its current Global Patient Safety Challenge, with a specific focus on improving poor processes and procedures.^10^ Their three categories for early priority action include high-risk situations (eg, hospitals and high alert medications), polypharmacy, and transitions of care. *We will address all three of these key categories in this project*.

### High Risk Medication Safety Situations Are Known

Transitions in Care, particularly in and out of hospital and High Alert Medications, are two primary high risk situations referenced by WHO.^10^ Even with evidence of under-detection, systematic reviews of the literature conclude that adverse drug events (ADEs) amongst older adults lead to approximately 1 in 10 hospitalizations.^11–13^ Canada-wide data suggest that adverse drug-related hospital admissions are directly correlated with the number of medications taken concurrently, the number of prescribers involved, possibly the number of pharmacies used, and to hospitalization within the previous year.^14^ In several studies, female sex was also a risk factor.

Once in hospital, patients (n=46,626) remain at high risk of ADEs, with a mean prevalence of 21.6% (SD 16.7,), 20.7% of these judged to be severe or life-threatening and 32.3% (SD 22.6%) judged to be preventable.^15^ In addition to the risk factors above, a complex patient (several comorbidities with several provider experts involved) is significantly more likely to suffer adverse events in hospital.^16, 17^ In Canada, medical and surgical services, because of the older age, high complexity and volume of patients seen, have the highest rates of adverse events overall.^16^

The period immediately following hospital discharge remains high risk with 37% of seniors sustaining medication-related harm (81% serious) within 8 weeks.^18^ In this systematic review, female sex was associated with medication-related harm.^18^ This leads to frequent readmissions and high costs. The outcomes of ADEs included prolonged length of stay, frequent readmission, emotional trauma, high costs, and death.^16, 19, 20^

### Polypharmacy and Potentially Inappropriate Prescribing (PIP) Are Important Signals

The term “polypharmacy” has multiple definitions, but the most common is the concurrent use of 5 or more medications daily by a single individual.^4^ The prevalence of polypharmacy in Canada is very high amongst seniors, at approximately 66%, with 27% of these taking 10 or more medications concurrently, a situation we label major polypharmacy.^4, 21^ While the number of medications used remains a useful signal, it is impossible to gauge the quality of medication regimens simply by the number of medications, as this does not account for the patient’s main diagnosis, comorbidities, risk factors, past history of medication use, or the benefit-harm ratio of the drug for that patient. For example, an older patient with diabetes frequently requires two glucose lowering medications, a statin for cholesterol, and two to three medications for blood pressure, just to manage their high cardiovascular risk without treating their other health problems. Thus, the medication safety target is <u>problematic</u> polypharmacy as opposed to appropriate polypharmacy.^22^

Medications which frequently lead to harms outweighing benefit in certain situations are termed ‘potentially inappropriate medications’ (PIMs). PIMs that have been associated with ADEs have been grouped together in medication screening lists, with the most evidence-based being the **STOPP** criteria.^23^ Randomized trials in Europe show that use of the STOPP criteria as a trigger for medication review for hospitalized seniors can improve the appropriateness of prescribing, reduce ADEs and reduce length of stay.^24, 25^ A cross-Canada study found that nearly 40% of seniors fill a prescription for at least one PIM per year, with the highest rates in women > 85 years of age and the most common PIM drug category being sedative-hypnotic drugs.^26^ The cost of these PIMS plus the cost of treating their adverse effects has been estimated to be more than $1.8 billion every year in Canada.^26^

### Interventions Should be Focused on Priority Areas

#### A. Priority Medications

Although STOPP is an excellent screening tool, there are too many alerts (80 in current iteration) to feasibly apply in hospitalized patients where timely discharge is a high priority.^25^ Analyses of Canadian and U.S. data on medication utilization and drug-related causes of hospitalization by our group and others, suggest recurring groups of very commonly, widely used medications as the main causes of drug-related hospitalizations.^14, 27, 28^ These are all medication families with proven benefit of varying clinical importance but also clinically important harm when not managed expertly. Thus, these are medications that should trigger review of the entire medication regimen to consider improvements. We have labelled these the high risk medications (HRM):

i. Anticoagulants,
ii. Analgesics including Opioids, NSAIDs and Glucocorticoids,
iii. Antimicrobials,
iv. Antineoplastic agents,
v. Glucose-lowering drugs for diabetes,
vi. Cardiac drugs including Diuretics and Digoxin,
vii. Sedative-hypnotics including Benzodiazepine receptor antagonists,
viii. Antipsychotic agents

Further details are included in Appendix 1. In many cases, the high-risk medication itself (e.g., anticoagulant, glucocorticoid, glucose lowering medication, antineoplastic agent, etc) is required for the patient but the dose may require adjustment, a tapering regimen might be appropriate, or a review for potentially serious drug interactions or medication burden identifies another medication that can be removed to decrease the patient’s overall risk of medication-related harm. We have previously developed an ‘Appropriateness of Prescribing Evaluation Questionnaire’ that has become the standard, holistic, medication appropriateness assessment tool.^29, 30^ Our ongoing development work suggests that while optimization of the high risk medications is feasible, it also opens up other opportunities for important improvements in medication management, including removal of medications and supplements with no benefit or with possibility of contamination, or substitution of more cost-effective alternatives.

#### B. Priority Patients

So-called high cost users (HCUs) have been an international priority target for quality and cost of care improvements for years.^31^ In a series of large population-based observational studies, we have shown that 5% of Ontarians generate 65% of the entire health care costs, approximately 70% of these are seniors (> 500,000), and these are complex patients with multiple hospital admissions, multiple providers, multiple diagnostic labels and problematic polypharmacy.^32–34^ Use of high-risk medications is very prevalent in this population and strongly predictive of future healthcare utilization and mortality in a dose- and duration-dependent manner compared to non-users.^32–35^

#### C. Priority Situations

Senior HCUs taking high-risk medications who are transitioning in, through, and out of hospital are at very high risk of serious ADEs. Hospitalization is a double-edged sword in that it defines high risk situations but also houses the expertise required to effectively intervene. This opportunity to optimize medication regimens for these inpatients is widely underutilized worldwide, due to a) huge pressures to just deal with the main problem requiring admission and get the patient discharged as quickly as possible due to bed shortages and b) lack of expertise amongst general medicine and surgery admitting services to complete an expert medication assessment quickly. In preparation for this trial, we recently completed a chart review of 100 randomly selected senior HCUs who were admitted to a Hamilton hospital (mean age 82 years), and found the mean rate of potentially inappropriate medications to be 2.8 per person. Only 16.6% of these had been addressed by the time of discharge.^36^

### Medication Safety Interventions Worth Evaluating

Two areas of innovation, heretofore not available for use in Canada, have matured sufficiently to be potentially effective, cost-effective, and highly feasible.

#### A. Innovation in Digital Health

The ability to date of digital health solutions, including electronic health records (EHRs) to meet the demands and complexity of modern medicine has been underwhelming.^37^ However, all patient safety leadership organizations including the Institute for Safe Medication Practices (ISMP), the Canadian Patient Safety Institute (CPSI), and the Agency for Health Care Research and Quality, agree that EHRs must be part of the medication safety solution, as system factors in addition to human factors, are always amongst the key root causes of adverse events. ^38–40^

<u>Epic</u> is a world-leading EHR software. It has won the Best in KLAS award for healthcare software suites 8 years in a row and is the EHR for the 20 top hospitals and the 20 top graduate schools for medical research in the United States.^41, 42^ St Joseph’s Healthcare Hamilton is the first large academic hospital system in Canada to go live (in 2017) with a build called Dovetale-Epic.^43^ This installation of Epic has led to rapid *innovation in both Clinical Care and in Research*.

*Clinical Care advances* relevant to this project include a) automated patient risk screening abilities, b) medication management alerts, c) improved physician-pharmacist communications, d) ability to easily maintain and connect an organized ‘circle of care’ for the patient (hospital and community providers, family, caregivers), and e) fully integrated telemedicine/telehealth compatible with a patient’s smart phone which facilitates and documents ‘virtual visit’ follow-up in the patient’s home. This technology extends the consultation reach of scarce, valuable specialties such as Clinical Pharmacology and Toxicology (CPT) to anywhere in Canada.

*Research innovation* includes novel major efficiencies in a) recruiting for trials, and b) quality and efficiency improvements in data collection and capture using secure transfer to industry-standard research data platforms (REDCap). Although a long time coming, EHRs have tremendous potential to improve the quality (richer data) while reducing the cost of RCTs.^44^ Since virtually all of the leading medical centres for clinical care and research in the United States use Epic, and several large additional hospital installations are imminent in Canada, this project will serve as a prototype pilot for a very large international research network serving a population of more than 150 million people.

#### B. Innovation in Clinical Expertise

Clinical Pharmacology and Toxicology (CPT) is one of the newest and smallest Royal College of Physicians - certified specialties in Canada. Its key mandate is improving the quality of drug therapy from bench to bedside to policy.^45^ Hamilton’s clinical pharmacologists who are also certified in Internal Medicine are unique in Canada in their ability and depth of experience in caring for complex elderly patients on multiple medications in need of evidence-informed review while in hospital and in follow-up. Their training and supervisory expertise has been recognized with multiple awards, and includes building capacity amongst trainees who will work elsewhere. In addition, the drug policy and formulary decision-making that is crucial to guiding cost-effective prescribing, is heavily influenced by CPT.

## METHODS

### Design

Pragmatic pilot randomized trial (RCT) at SJHH with 1:1 patient-level concealed-allocation randomization with blinded outcome assessment and data analysis. Randomization provides the highest quality methods to minimize bias, the pragmatic design ensures relevance to clinical practice essential for implementation, and a pilot RCT addresses feasibility of a large definitive subsequent RCT directly without waste of research dollars.^46, 47^ A study flow diagram is shown below in Figure 1.

**Figure 1.**
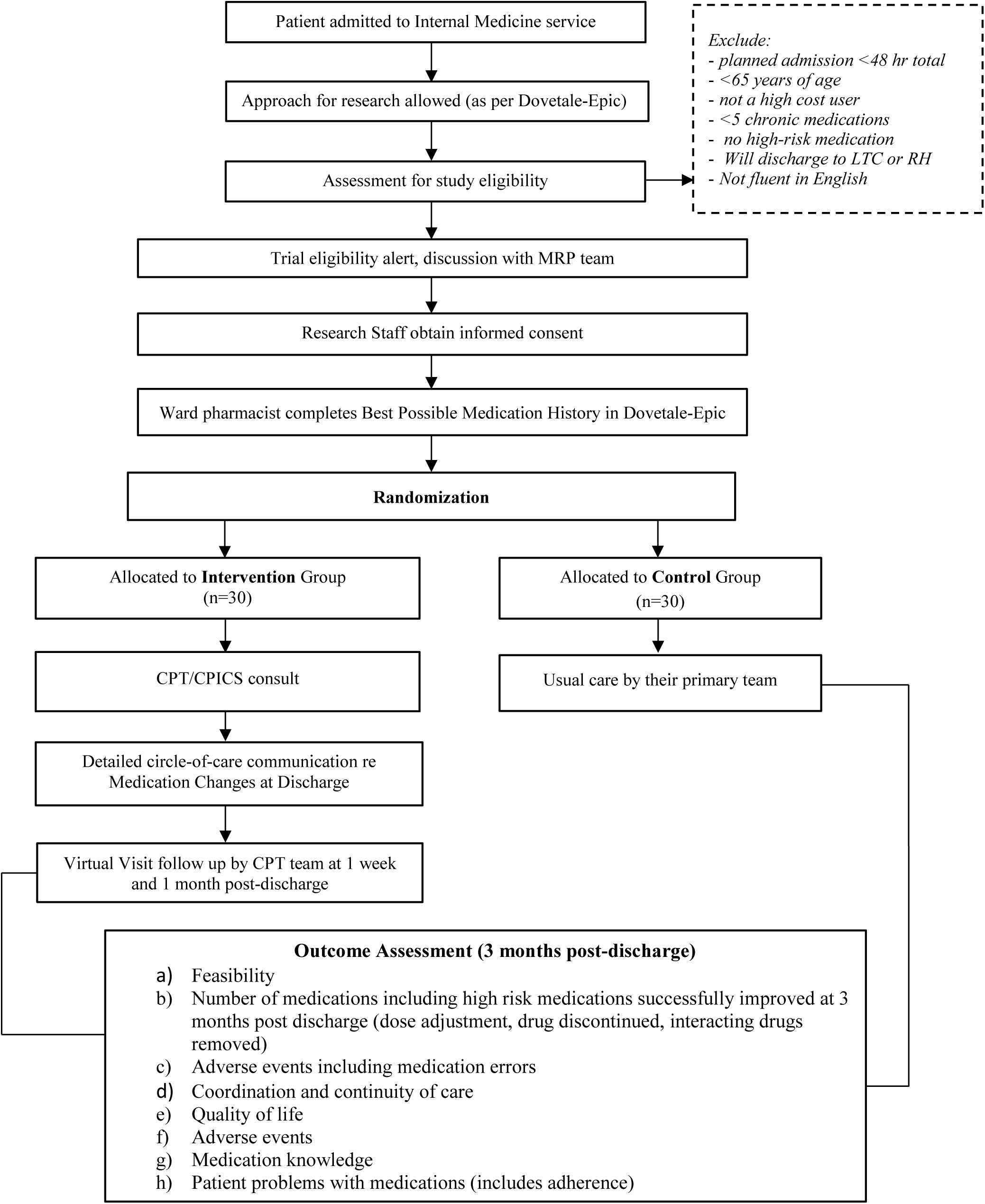
IMPROVE-IT HRM RCT Flow Diagram

### Participants

Adults over the age of 65 years who are admitted to Medicine services for more than 2 days, who are high cost users defined as at least one other hospitalization within the previous year, who are taking 5 or more chronic medications including at least one high risk medication and provide informed consent. It is estimated that at least ten patients daily meet these eligibility criteria.

### Recruitment and Randomization

The Research Institute of St Joes Hamilton recently approved a policy that every patient admitted can be approached to participate in studies unless they decline (Access Research). Dovetale-Epic tracks this ‘may approach/do not approach’ status for each patient in real time. This effectively alleviates a barrier as the (usual) necessity of relying on the patient’s core team to refer a potentially eligible patient is a major barrier to recruitment, simply due to distraction of the team with the clinical priorities.

We are working with the Dovetale team to have all potentially eligible patients screened for age, HCU status (defined as at least one previous admission in the prior year) and the presence of five or more chronic (not prn) medications with one or more high risk medication. Screen eligibility-positive patients will be posted to the current Clinical Pharmacology and Toxicology team’s intake list. Once eligibility is confirmed, the ward pharmacist will ensure that an accurate BPMH (best possible medication history) is completed and documented appropriately in Dovetale-Epic. An accurate BPMH is critical for an accurate discharge medication reconciliation. Research staff will check with the most responsible physician (MRP) team regarding eligibility (this stage picks up on delirium and dementia, for example). The research team will then approach the patient to invite participation, introduce the study and complete the informed consent process which includes answering questions, completing a short Capacity to Consent questionnaire (Appendix 2), and Informed Consent document.^48^

Patients with cognitive impairment will not be excluded as they constitute a vulnerable group in need of assistance, but they must have a primary caregiver who assists them with medications and who is willing to sign informed consent if the patient fails the Capacity to Consent questionnaire.^48^ For this pilot trial, either the patient or caregiver must be fluent in English.

Patients will be randomly allocated to the intervention or control arms in a 1:1 ratio in permuted blocks using a statistician-formulated randomization schedule that will be available online 24/7.

A validated patient frailty index will be incorporated at baseline as a prognostic marker^49^.

### Intervention

Our intervention follows the general Innovative Practices framework recommended by Health Quality Ontario for Transitions between Hospital and Home, and uses the specific approach in Figure 2.^50^

**Figure 2.**
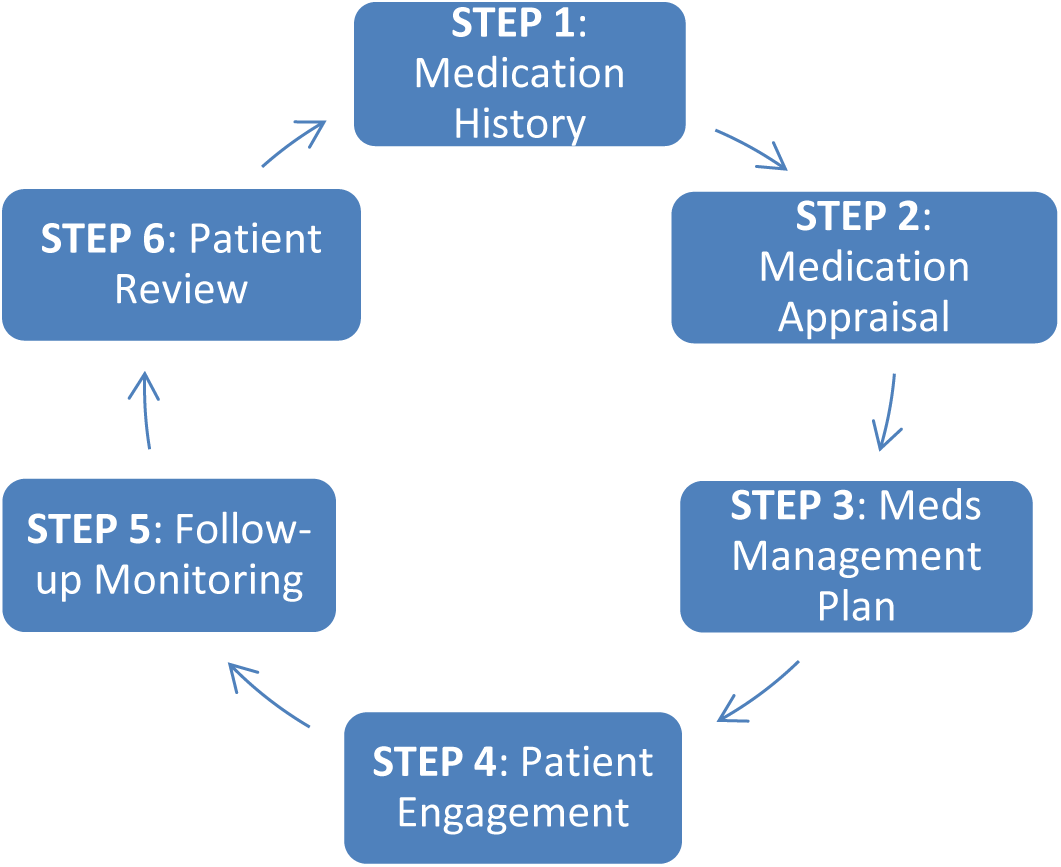
IMPROVE-IT HRM Clinical Approach

#### CPICS (Clinical Pharmacology Inpatient Consult Service) Notification

Randomization to the intervention arm will trigger a request for a CPT (Clinical Pharmacology & Toxicology) consult, using our Clinical Pharmacology Inpatient Consult Service (CPICS) workflow. The CPT team consists of a unique combination of expert Clinical Pharmacologists (Royal College physician specialist in patient assessment and diagnosis, drug therapy, adverse drug events, drug access, safe prescribing, cost-effective medication management) with additional expertise in evidence-based geriatric prescribing, multi-morbidity, and de-prescribing; medicine and pharmacy trainees; and a research assistant. There are fewer than 20 such CPT specialists in Canada, four of them located at SJHH with the largest cumulated experience of any medical centre at more than 90 physician-years.

#### Initial Consult

The initial CPT consult for each patient includes a comprehensive patient assessment including demographics, social situation, drug coverage insurance, functional status (activities of daily living, instrumental activities of daily living), cognition, frailty markers, level of caregiver involvement, past medical history and current problems, allergies and intolerances, detailed medication history including reminder aids and methods of accessing medication, physical exam and review of current and historical laboratory and diagnostic imaging results. These details are structured data items in EPIC behind a customized CPT consult, progress or discharge note structured template, which ensures consistency of intervention across the four consultants.

In addition, the CPT team will document a detailed ‘circle of care’ for each patient (their primary caregiver, hospital MRP team, family doctor, community physician specialists, community pharmacists and home care), and will identify all potential high-risk medications targets that the patient is taking or is due to resume post-hospitalization. Using patient preference elicitation methods and motivational interviewing, priorities for medication optimization will be negotiated.^51–56^ Short patient infographics endorsed by the Canadian Medication Appropriateness and Deprescribing Network (CADeN) will be used as educational materials.

#### CPICS Follow-up

While the patient is still hospitalized, the high-risk medications care plan begins and the team ensures close communication with the MRP (Most Responsible Physician) admitting team, coordinates a detailed discharge medication reconciliation (documenting medication changes with rationale, formulating an accurate discharge prescription including rapid access to new medications), ensures circle-of-care communication and sees the patient via Virtual Visit twice in follow-up at 1 week and 1 month after hospital discharge to complete the care plan.

### Control

These patients will receive usual care by their primary team. This means that the MRP team is responsible for coordinating medication management at discharge and post-hospital follow-up, as is currently practiced.

### Outcomes

The Core Outcome Set for Interventions to Improve Polypharmacy in Older People was consulted to assist with our selection of outcomes.^57^ Core outcome sets are consensus-based guidelines from groups of clinicians, patients and methodologists on which outcomes with which metrics, are the most important to be measured in prospective studies.^58^ In addition, we consulted other polypharmacy/deprescribing trials for recommended patient-important outcomes.^59–64^

For this pilot RCT, we will analyze outcomes according to a set of Primary and Secondary Outcomes. (see details in Table 1 and Table 2.)

**Table 1.** IMPROVE-IT HRM OUTCOMES^29, 30, 57, 59, 60, 65, 68, 91, 92^.

|  | **Outcome** |
| --- | --- |
| Feasibility | **1.** Participant recruitment rates |
|  | **2.** Participant retention rate |
|  | **3.** Trial resource utilization, costs |
|  | **4.** Recommendation acceptance rates by MRP team |
|  | **5.** Recommendation acceptance rates by patient |
|  | **6.** Recommendation adherence rates by patient |
|  | **7.** CPICS consultation volume capacity |
|  | **8.** Potential to intervene entirely virtually (online) |
| Clinical | **1.** Medication Management Outcomes<br>a) # drug therapy problems improved<br>b) # High Risk medication problems improved<br>c) # PIMs tapered or discontinued (using APEQ)<br>d) # medications per patient end-study versus baseline |
|  | **2.** Adverse drug events including adverse drug withdrawal event |
|  | **3.** Medication Errors, including preventability |
|  | **4.** Patient Problems with Medications Questionnaire (includes general adherence) |
|  | **5.** Coordination and continuity of care |
|  | **6.** Patient Quality of Life (EQ-5D-5L and Medication-related QOL) |
|  | **7.** Patient Knowledge of Medications |
|  | **8.** Satisfaction with Care (Patients and Providers) |
|  | **9.** Health Resource Utilization, including Clinical events of death, ED visits, hospitalization or unplanned physician visits; medication costs, to estimate cost - effectiveness |

\* An Adverse drug event (ADE) is defined as “Harm caused by exposure to a drug” whereas an adverse drug reaction (ADR) is a subset of these events, where harm is caused by a drug under appropriate use (i.e. at usual doses) – adjudicated using Leape and Bates scale: 6- definitely due to medication, 5- probably due to medication, 4- possibly due to medication, 3- possibly due to disease, 2- probably due to disease, 1- definitely due to disease (5 and 6 considered ADE).<sup>66</sup> Medication Errors have been defined as an error (of commission or omission) at any step along the pathway that begins with prescribing and ends with the patient taking the medication.<sup>66</sup>

**Table 2.**
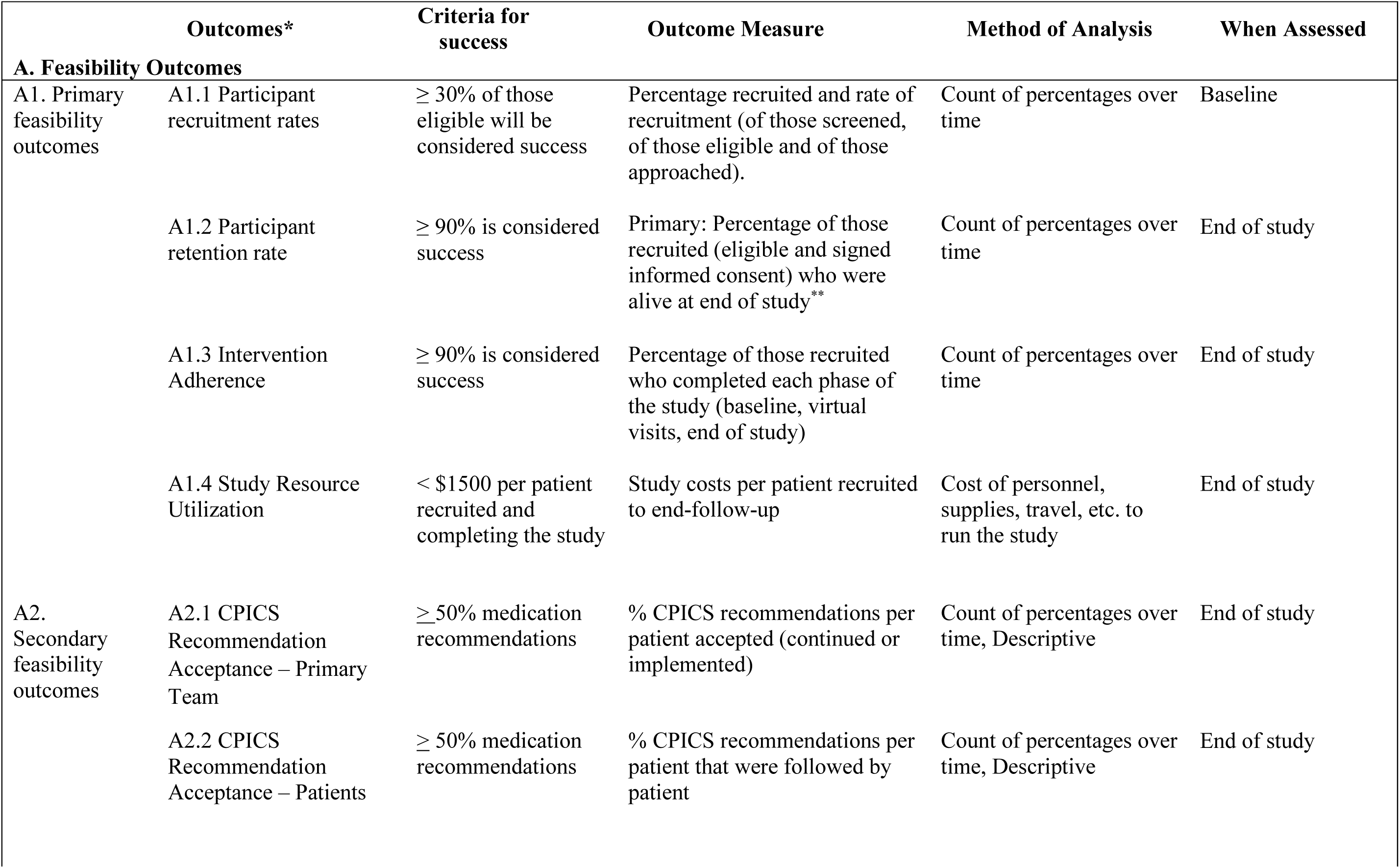
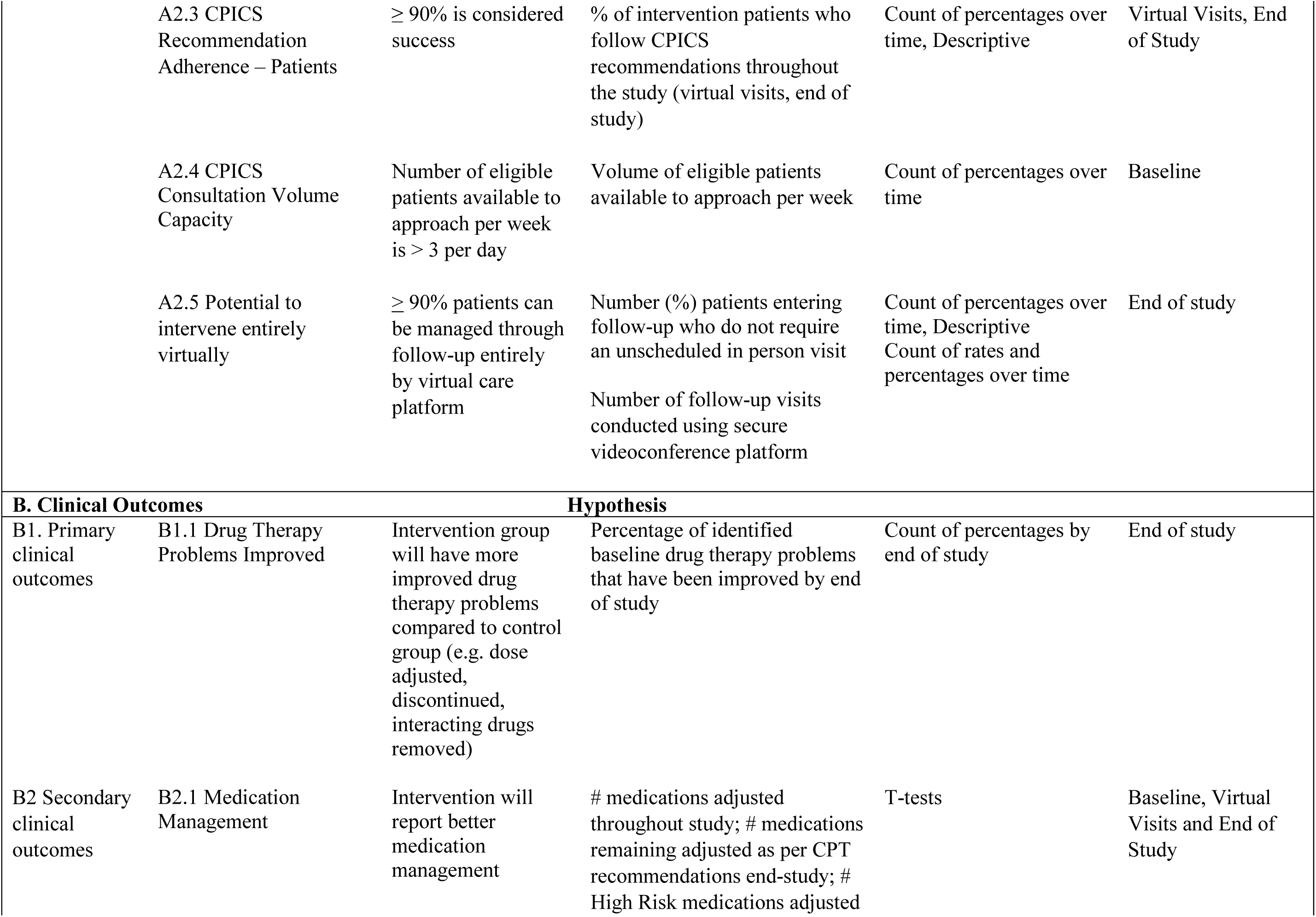
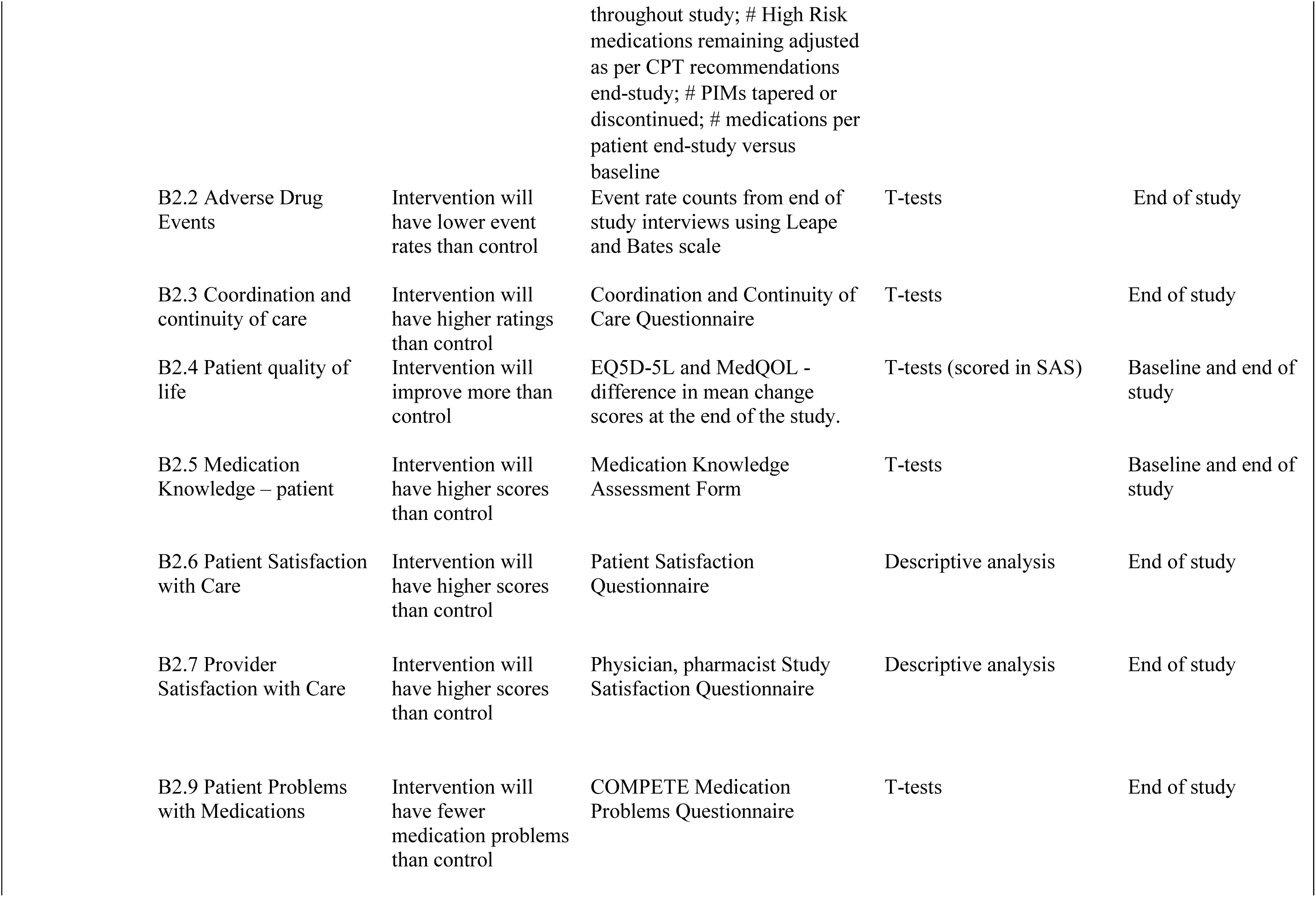
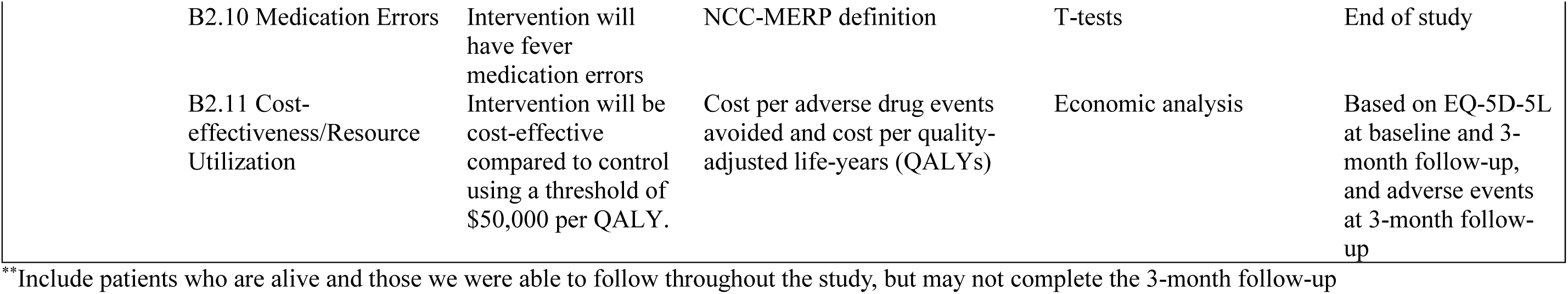
IMPROVE-IT HRM Study Outcomes and Measures.

### Primary Outcomes

There are two types of primary outcomes that will be evaluated:

a. The *Primary <u>Feasibility</u> Outcomes* will be recruitment and retention rate for eligible patients, and the estimated resources required per patient to complete the main trial. We aim for at least 30% recruitment of those eligible, 90% retention of those recruited, and no more than $1500 per patient spent on running the pilot trial.
b. The *Primary <u>Clinical</u> Outcome* will be the number of drug therapy problems improved including the number of high-risk medications improved (for example, dose adjustment, discontinued, seriously interacting drugs removed) at 3 months post-hospital discharge end-study visit.

### Secondary Outcomes

a. The *Secondary Feasibility Outcomes* of CPT consultation include: CPICS recommendation acceptance by primary team and by patients, consult volume capacity, and potential to apply the intervention entirely through virtual visits.
b. The Secondary Clinical/Patient-important outcomes will include:

1. Other medication management outcomes (details in Table 1)
2. Adverse drug events using the standard definition of harm caused by use or inappropriate use of a drug, and including rating of preventability using the ‘best practice’-based definition of Woo et al – inappropriate drug or dosage or route or frequency of a drug given the patient’s age, diagnoses, weight, organ function; or administration of a drug for which the patient has a major allergy or intolerance; or inappropriate laboratory monitoring.^65^ Specific rating of ADE will use the standard Leape and Bates scale: 6-definitely due to medication, 5-probably due to medication, 4-possibly due to medication, 3-possibly due to disease, 2-probably due to disease, 1-definitely due to disease.^66^
3. Medication errors – characterized as errors in prescription, dispensing, or administration using standard NCC-MERP definitions and classification.^67^ A Medication Error is an error (of commission or omission) at any step along the pathway that begins with prescribing and ends with the patient taking or not taking the medication. ^68^
4. Patient Problems with Medications: The COMPETE Medication Problems Questionnaire measures problems with medication access, handling, beliefs and adherence.^69^
5. Medication Knowledge Assessment: this will be assessed using the Medication Knowledge Assessment form, which tests knowledge of medication name, indication, dosage instructions, and precautions.^70^
6. Coordination and Continuity of Care: Adapted from Health Quality Ontario’s draft guidance and a Rand instrument, the Coordination and Continuity of Care Questionnaire is designed to measure the quality of the transitional and follow-up care. We will focus on medication reconciliations and education, and circle of care communications.^71, 72^
7. Patient Quality of Life: The EQ-5D-5L is the 5-level classification system of the EQ-5D, a measure of health status from the EuroQol Group, which also contributes incremental utility values to the cost-effectiveness evaluation.^73^ Using EQ-5D-5L, respondents are asked a short series of questions about mobility, self-care, usual activities, pain discomfort, and anxiety/ depression, as well as a summary visual analogue scale. This scale, which provides utility measurements, has been well validated for the Canadian population.^73–75^ We will also use a ‘condition-specific’ QOL measure, the Medication-related Quality of Life measure which is designed for people with polypharmacy.^76^
8. Satisfaction with Care: Satisfaction reported by patients and by key health professionals is one of the recommended outcomes to report in medical research, as it may influence adherence.^77, 78^ This outcome will be assessed by the Patient/Caregiver Study Satisfaction Survey and by the Provider Study Satisfaction Survey.^79^
9. Health Resource Utilization: This is a key outcome to determine cost-effectiveness and cost-utility which then determines whether health care systems might pay for this type of care.^80, 81^ To improve accuracy and feasibility, we will estimate costs based on healthcare utilization, concentrating on Emergency Department visits, hospitalizations, unplanned physician visits and medication costs, including out-of-pocket medication costs in follow-up.^72, 81^. The analysis will follow the guidance of international and Canadian guidelines.^82, 83^

Outcomes will be collected through a mix of patient interview and chart review. The impact of sex, gender, age, social support, socioeconomic status, cognition, number of medications, and location of discharge, will be examined as potential interaction variables with the intervention and as predictors of outcomes. Sensitivity analyses will assist with determination of potential for cost-effectiveness of the intervention overall and in selected subgroups.

Geographic wards in the hospital allow for evaluation of the intra-cluster correlation (ICC), which based on past experience, we expect to be low. If ICC proves to be high, more complex cluster RCTs will be required. Additionally, barriers and facilitators to success of the primary outcomes, in terms of process and management issues will be evaluated. These will be used to determine whether a large definitive research study is likely to be feasible, taking into account the practical aspects of managing and funding the project.

### Follow-up

Patients will be followed until three months post-hospital discharge or until death, whichever occurs first. The challenges and uncertainties posed by the COVID-19 pandemic have highlighted the role and importance of telemedicine, which this trial will utilize for patient follow-up visits. These visits will be conducted by videoconference via Dovetale-Epic or Ontario Telehealth Network, or by phone call, depending on the patient’s digital technology capability.

### Sample size

Our calculations, based on two independent study groups, measuring a continuous primary outcome, allowing a probability of a Type 1 error of 5%, suggest that to have 80% power to determine an improvement in number of drug therapy problems improved from 3 (Standard deviation 1.4) to 2.5, would require a total sample size of 248 or 124 per group.^84^. Since this is a pilot RCT, we will aim for 30 patients per group or 60 in total as this number is likely to provide adequate evidence regarding feasibility for ramp-up and actual sample size.

### Allocation

Participants who meet all the inclusion/exclusion criteria at screening and have completed informed consent will be enrolled in the study, complete baseline assessments, then will be randomized via a computer-generated randomization sequence to one of the two study arms, intervention or control.

A statistician will prepare the randomization schedule.^85^ The pilot is being carried out at a single site, although an eventual trial will be multicentre.^86^ To restrict treatment group imbalance a maximal tolerable imbalance between treatment groups will be incorporated into the schedule.^87^ The randomization schedule will be produced by a program written in R and implemented in REDCap where each patient’s treatment assignment will be available on-line to the research staff only at the time of randomization. This process ensures allocation concealment, and randomization awareness where necessary, for example for the intervention staff.

### Blinding

Since this is a pragmatic RCT, it will not be possible to completely blind patients or their providers, however outcome data collectors, adjudicators and statisticians will be blinded to group allocation until analysis is completed at the end of the study.

### Data Collection Methods

Trained research staff will conduct the interviews with the patients or caregivers, entering data electronically on study laptops directly into REDCap case report forms. The participants’ medical records will be reviewed to abstract data on baseline characteristics, medical history, and medication information. Strategies to promote participant retention and complete follow-up include reminding participants in advance of their end-of-study visit and communicating by email if email address is provided at baseline. Participants who drop out of the study will have their data to that point retained in the study, as approved by REB, to avoid bias. The reasons for study non-completion will be recorded.

### Data Management

REDCap (Research Electronic Data Capture) is our study software platform – secure, web-based, providing interfaces for validated data capture, role-specific access, audit trails for tracking data manipulation and exports, automated export procedures to SAS and encrypted transmissions.^88, 89^ Paper study documents including signed informed consent forms will be stored in our secure research office once they are scanned into REDCap study files. Regular data quality checks, such as automatic range checks, will be performed by the study team to identify data that appear inconsistent, incomplete, or inaccurate.

Patients are not identifiable in the project results database. The identifying information required for the clinical team to deliver the intervention is kept in a separate database. Access to the final dataset will be restricted to the core research team.

### Statistical Analyses

The reporting of the results of this trial will follow the CONSORT extension for pilot trials.^90^ We will use descriptive statistics for presentation of baseline variables and adequacy of follow-up. Feasibility analysis including recruitment rate (≥30% is considered success), participant retention rate (≥90% to end of study is considered success), study resource utilization required (less than CAD $1500 per patient recruited), management assessment, and scientific assessment, will be descriptive.

Analysis will use intention-to-treat methods with censoring only if the patient dies or drops out of the study with refusal of negotiated further assessments. A sensitivity analysis of the subgroup of patients who received all planned follow-up intervention calls and completed the end-study data collection (per protocol analysis), will be carried out. Research staff and statisticians will review outcome data and analysis blinded to group identification.

The primary clinical endpoint will be number of drug therapy problems improved including the number of high-risk medications improved, as measured by chi-squared and t-tests. Secondary endpoint analyses of the coordination and continuity of care, patient quality of life, medication knowledge, satisfaction with care (providers & patients), and resource utilization, will be analyzed using t-tests. The incidence of the adjudicated individual clinical events (all-cause hospitalizations and emergency department visits) will be analysed using the methods described above for drug therapy problems. Public unit costs from Ontario will be used to cost healthcare resource utilization collected as part of the trial. Using an area under the curve approach, quality adjusted life years (QALYs) will be determined by weighting the EQ-5D-5L health utility scores by time spent in health state. Costs and outcomes (i.e., QALYs, adverse clinical events) between the groups will be compared from a public payer perspective.

Given the short follow-up, low risk of the trial and pilot design, no interim analysis or imputation for missing data is planned. All statistical analyses will be performed using SAS V9.4 software (SAS Institute Inc., Cary, NC, USA).

### Data Monitoring

Any serious adverse event will be reviewed by our Trial Steering Committee (TSC) within a week of detection, to discern any attribution to our procedures. If found to be due to our coordination procedures, the trial steering committee will recommend whether modifications are indicated. The TSC will be composed of individuals with expertise in clinical trials, chaired by the lead statistician, include the PI, the operational statistician plus a methodologist independent of the study team. Similarly, since this is a short pilot pragmatic RCT where no harm is expected and adjustment of trial procedures may be necessary for feasibility, no formal external auditing of trial conduct is planned. There is no requirement for additional ancillary and post-trial care for those who might come to harm while in the trial, as usual medical care which covers this eventuality is already in place.

### Privacy and Confidentiality

Study data will be entered into REDCap, which is a secure database, and access to REDCap will only be provided to approved research staff who are trained in general research guidelines (i.e. GCP, TCPS2, privacy tutorial). Patient identifiers will be stored in a separare REDCap folder with limited access. All data on the REDCap server is encrypted and all access is audited. A paper copy of the link between study ID and patient identifiers will be stored in a locked cabinet in a locked cabinet inside our locked research offices in the hospital, which is accessed only by our research staff. This link, both paper and electronic, will be destroyed at the end of the study. Any other hard copies of study related information (i.e. signed Informed Consent Forms), will be transported within SJHH by the research staff and stored in our secured offices. Patients will be assigned a study ID that will be recorded on their CRFs and no direct identifiers will be used on the Case Report Forms.

## IMPACT OF RESEARCH

By using an integrated KT approach with clinicians, patients, researchers, drug policy advisors, information technology advisors, quality improvement advisors, and hospital and national medication safety administrators involved in the project from protocol to dissemination, we hope to strengthen and broaden the usual dissemination of research to practice and policy. However, this is a pilot study, so the main dissemination question will revolve around whether there is sufficient value to justify a larger, definitive clinical trial. Given the major improvements in research efficiency and international research product dissemination that EPIC affords us, we will be cultivating a Canadian EPIC-based research network amongst the impending participating hospitals.

## ETHICS

The study has been approved (study #7598) by Hamilton Integrated Research Ethics Board. Significant protocol modifications will be proactively communicated to the research ethics boards through study amendments to obtain approval prior to the changes being implemented. Each modification will be assessed to determine whether it warrants communication with trial participants.

## Data Availability

All data produced in the present study are available upon reasonable request to the authors.

---

**Appendix 1.**
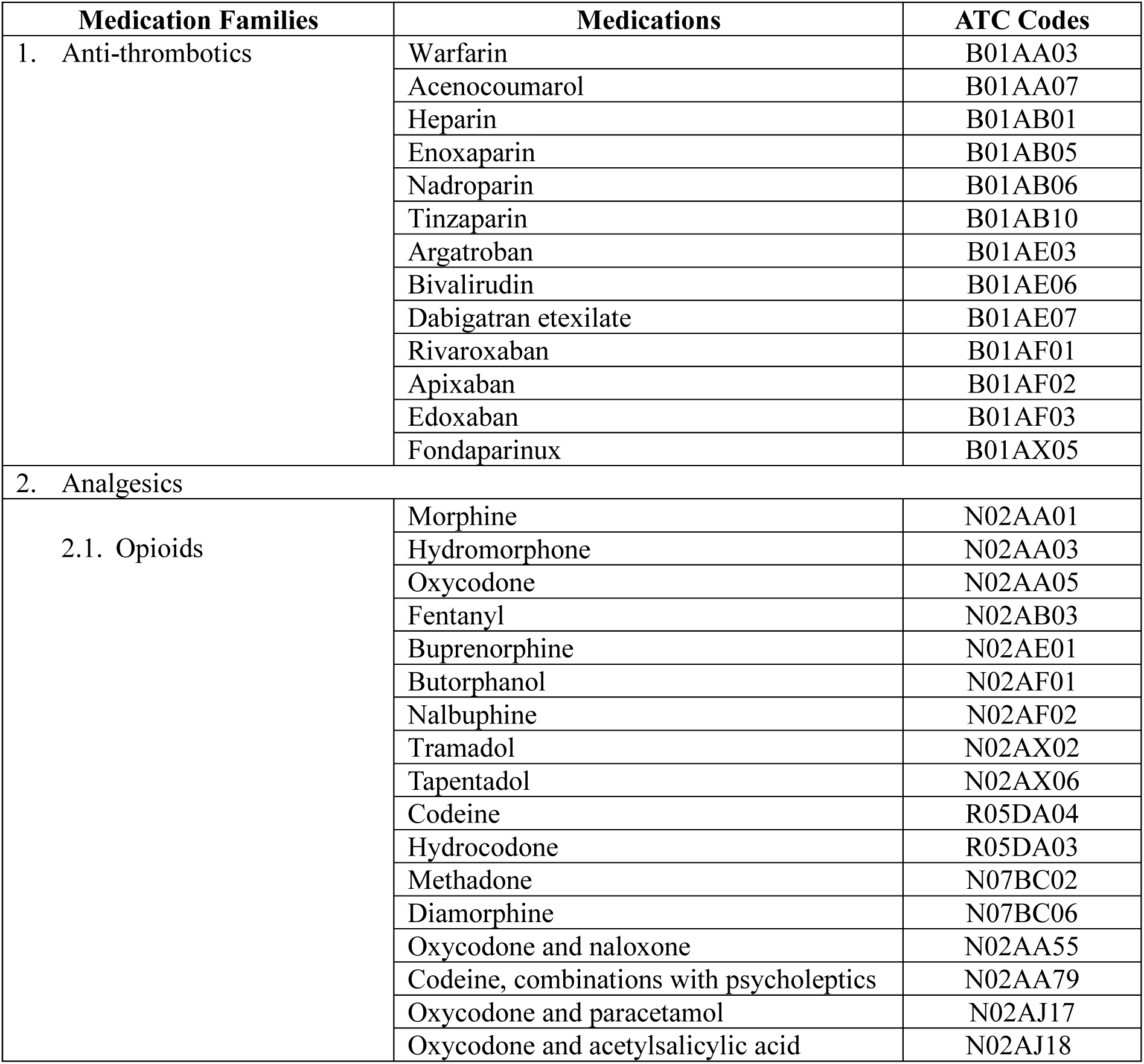
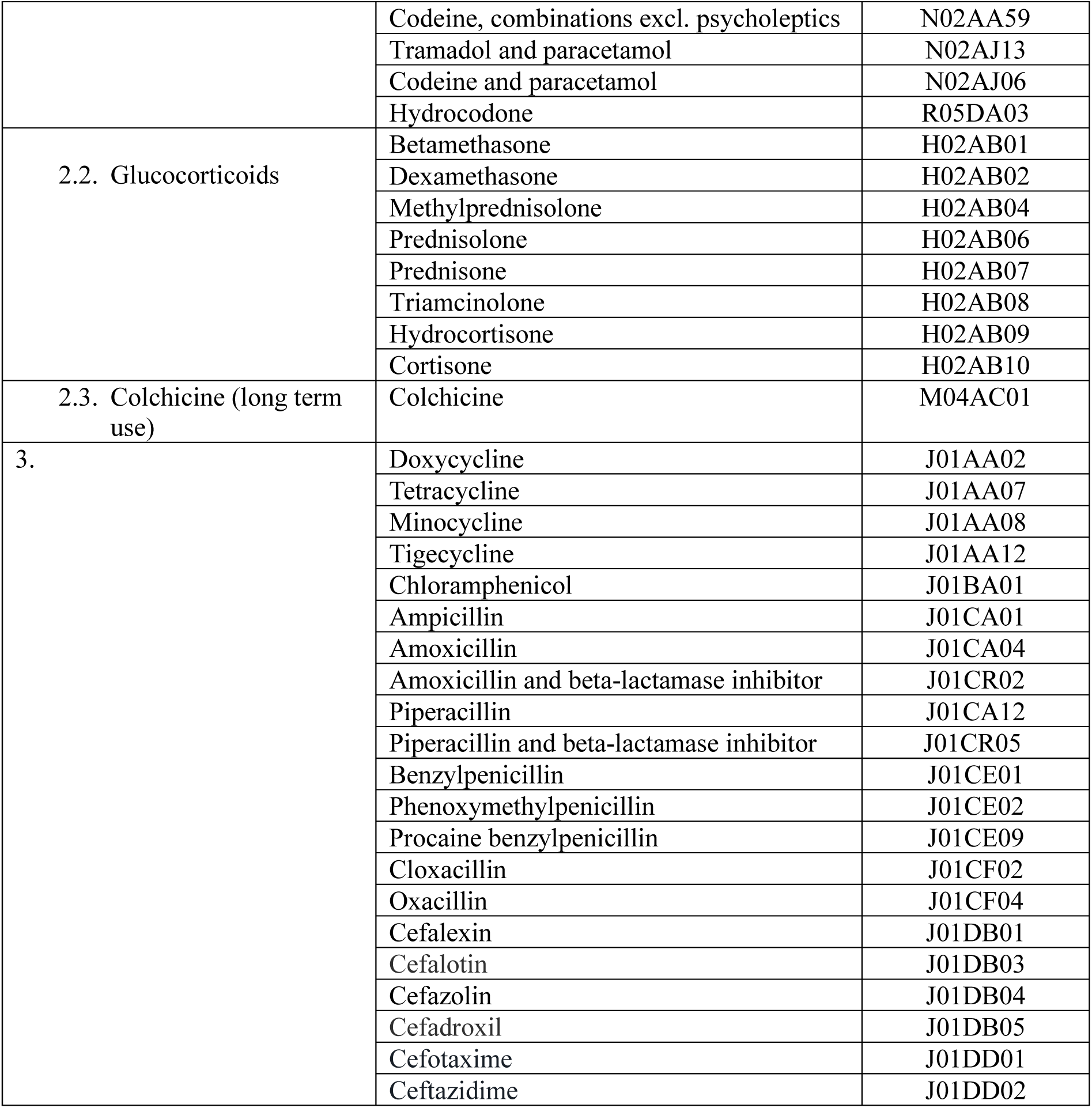
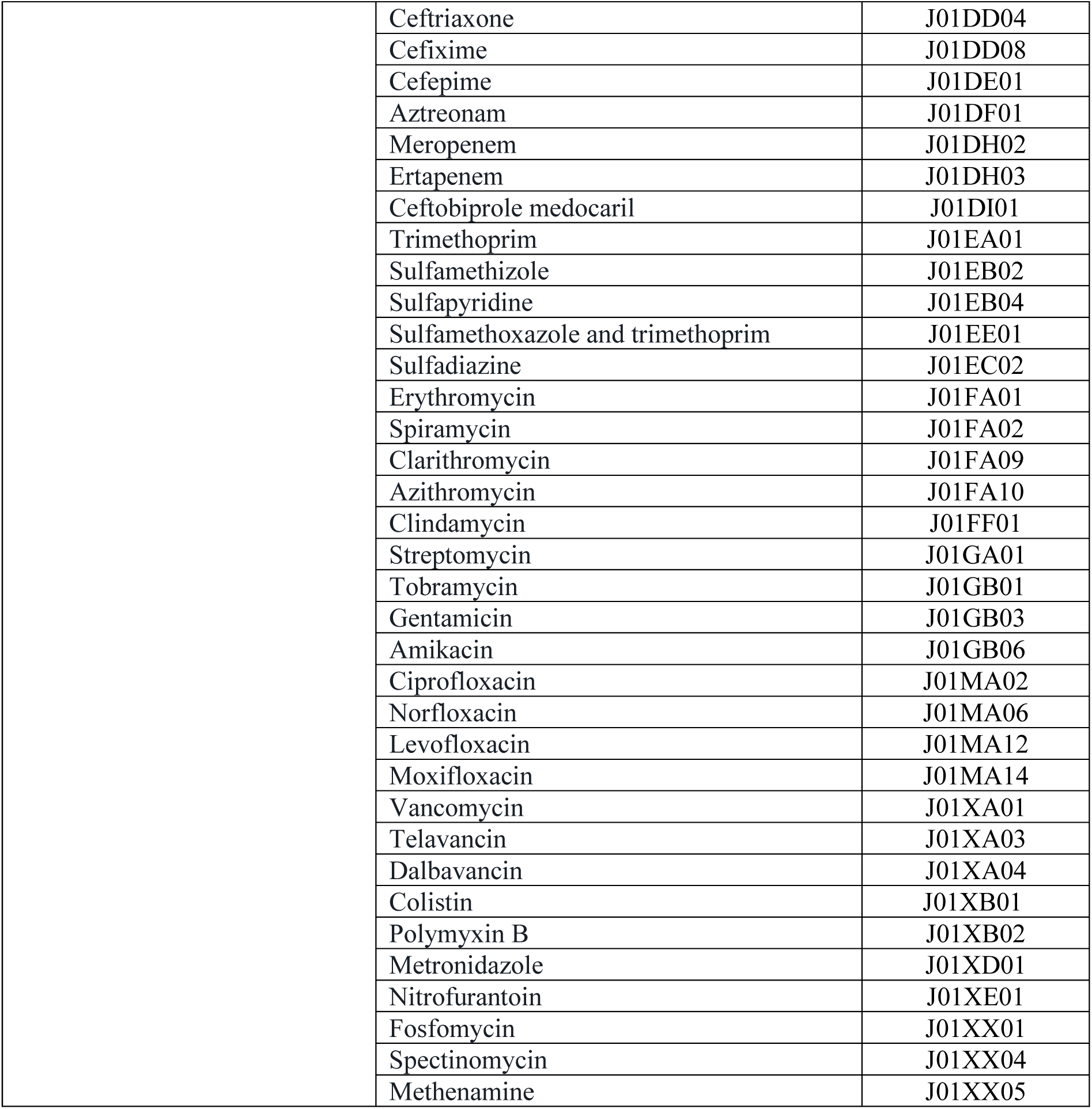
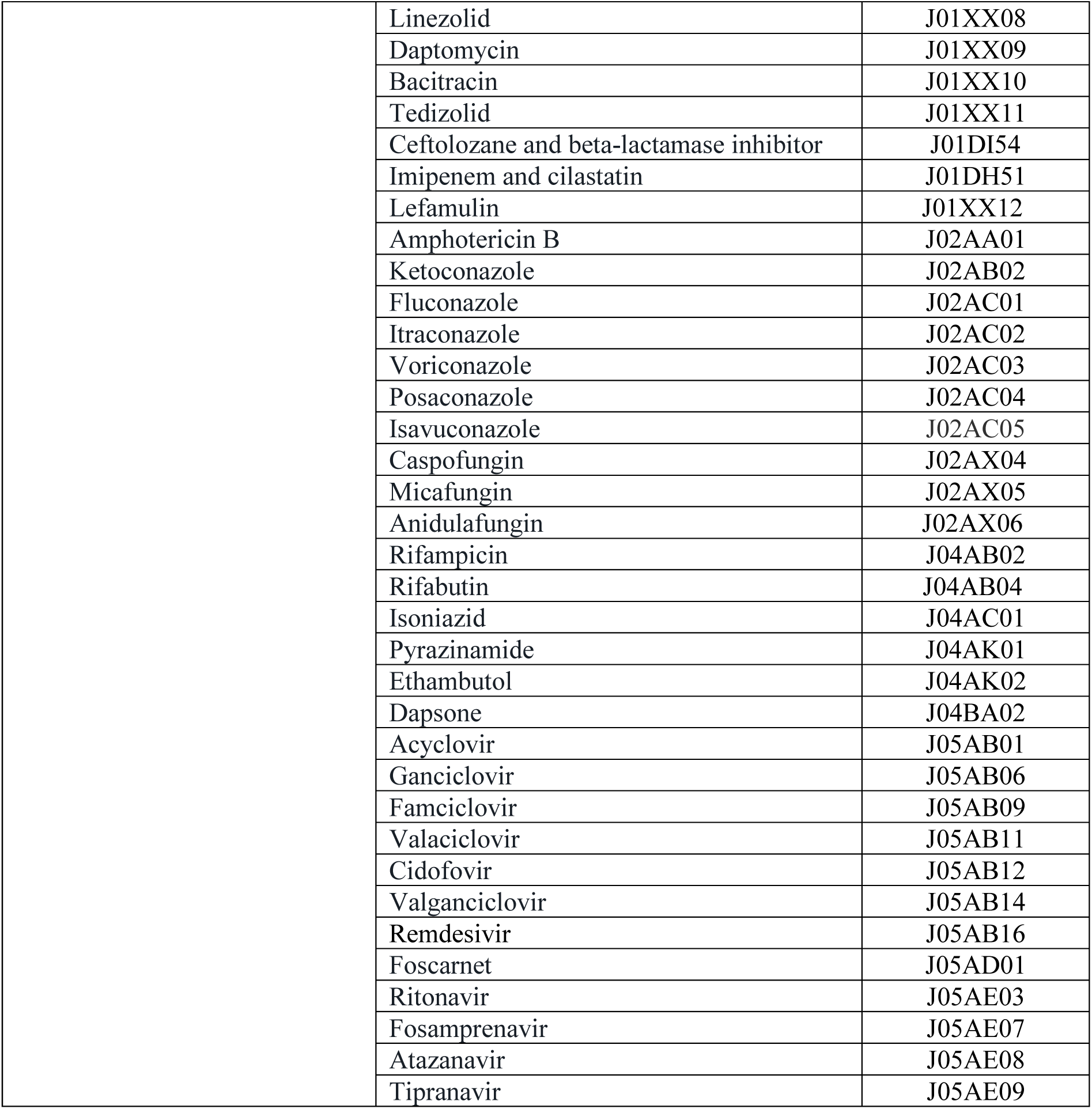
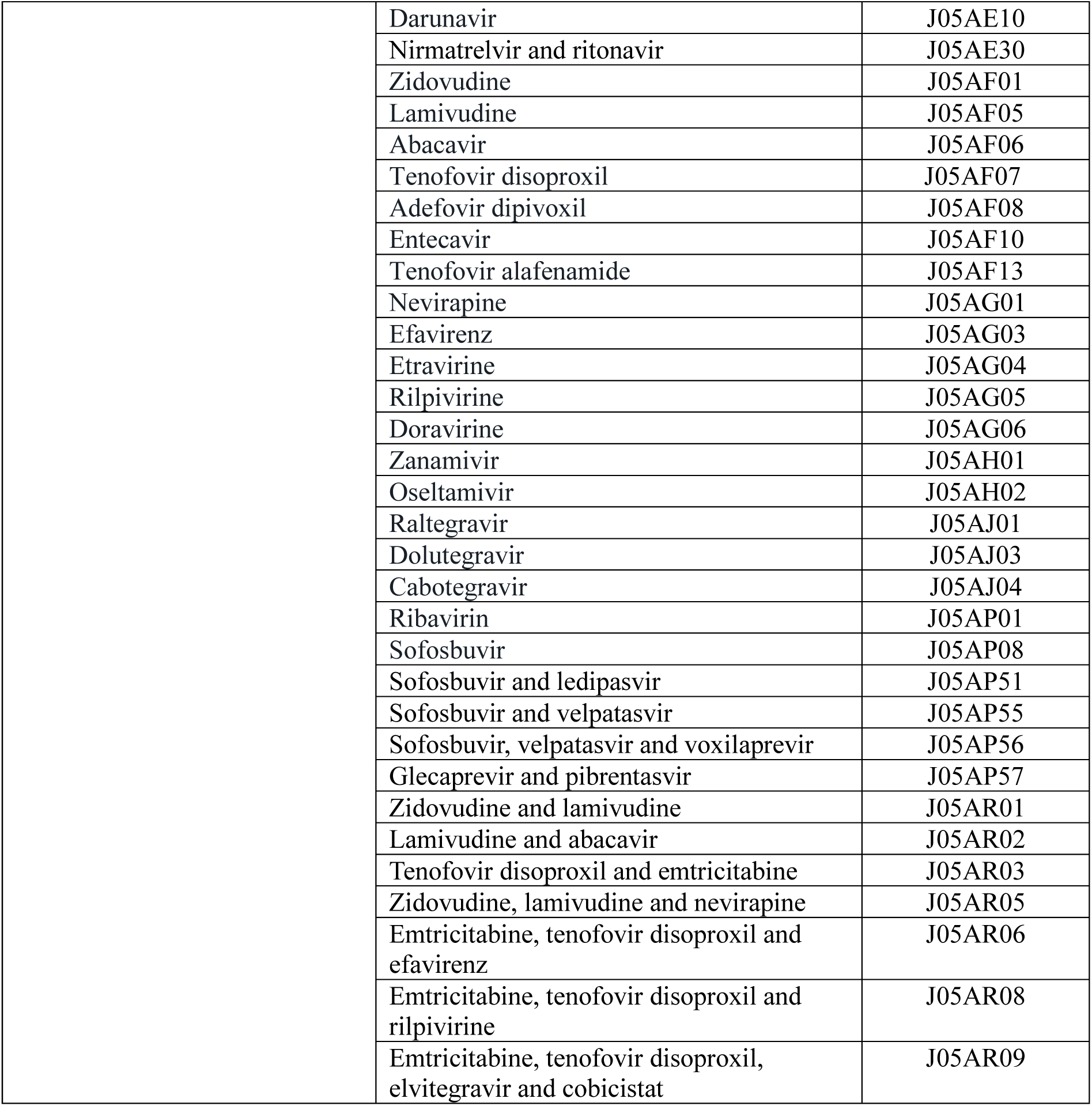
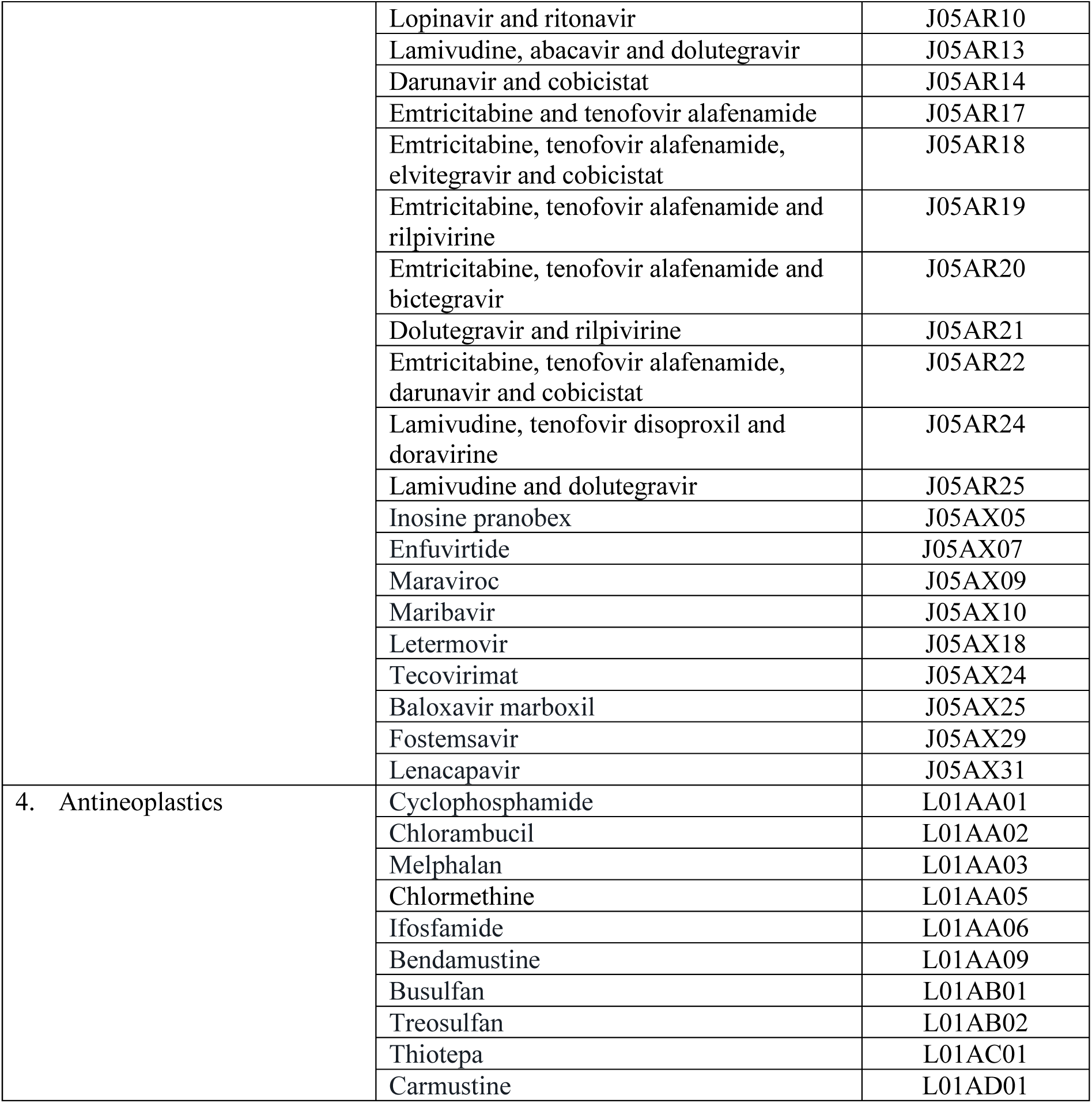
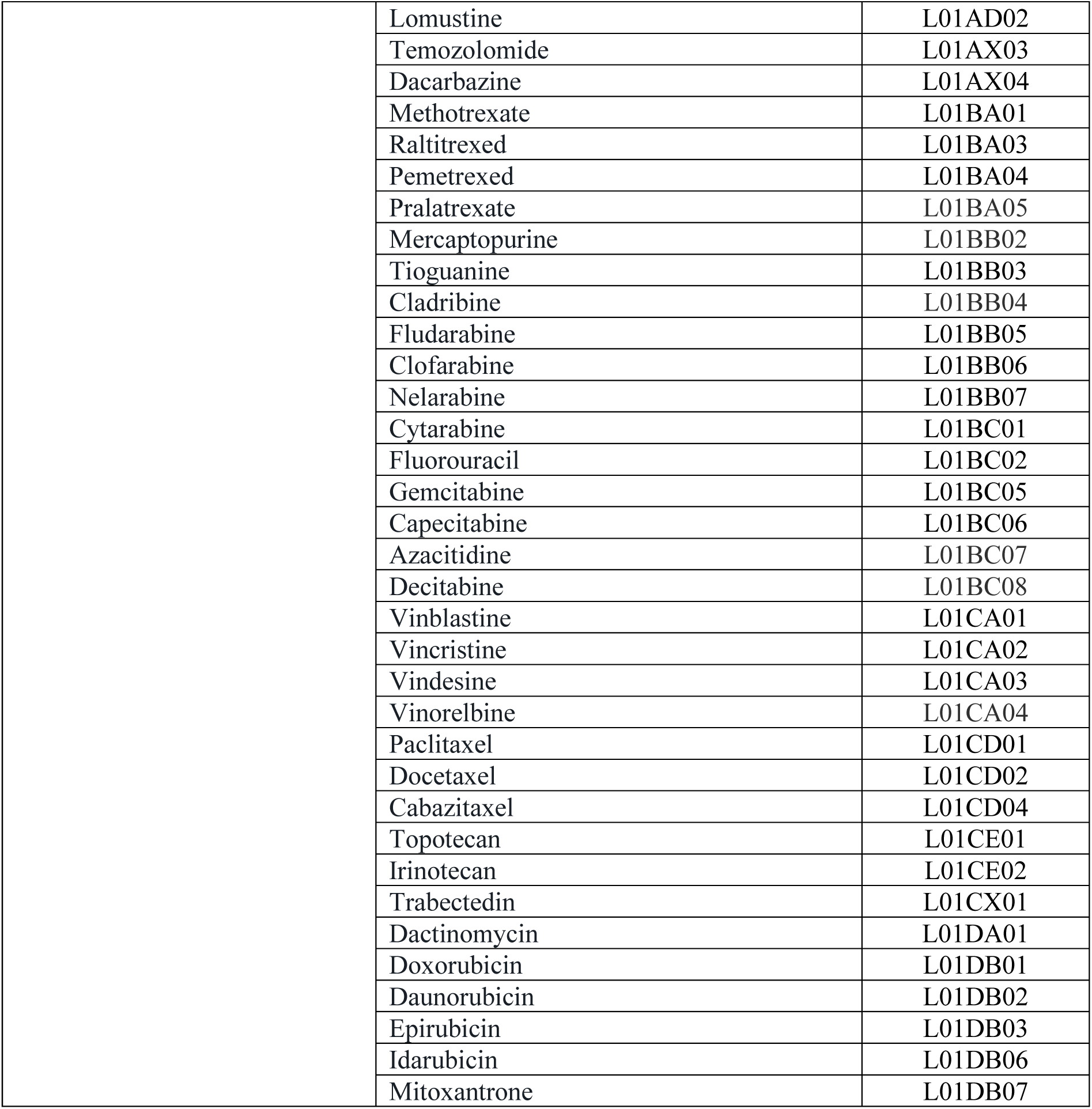
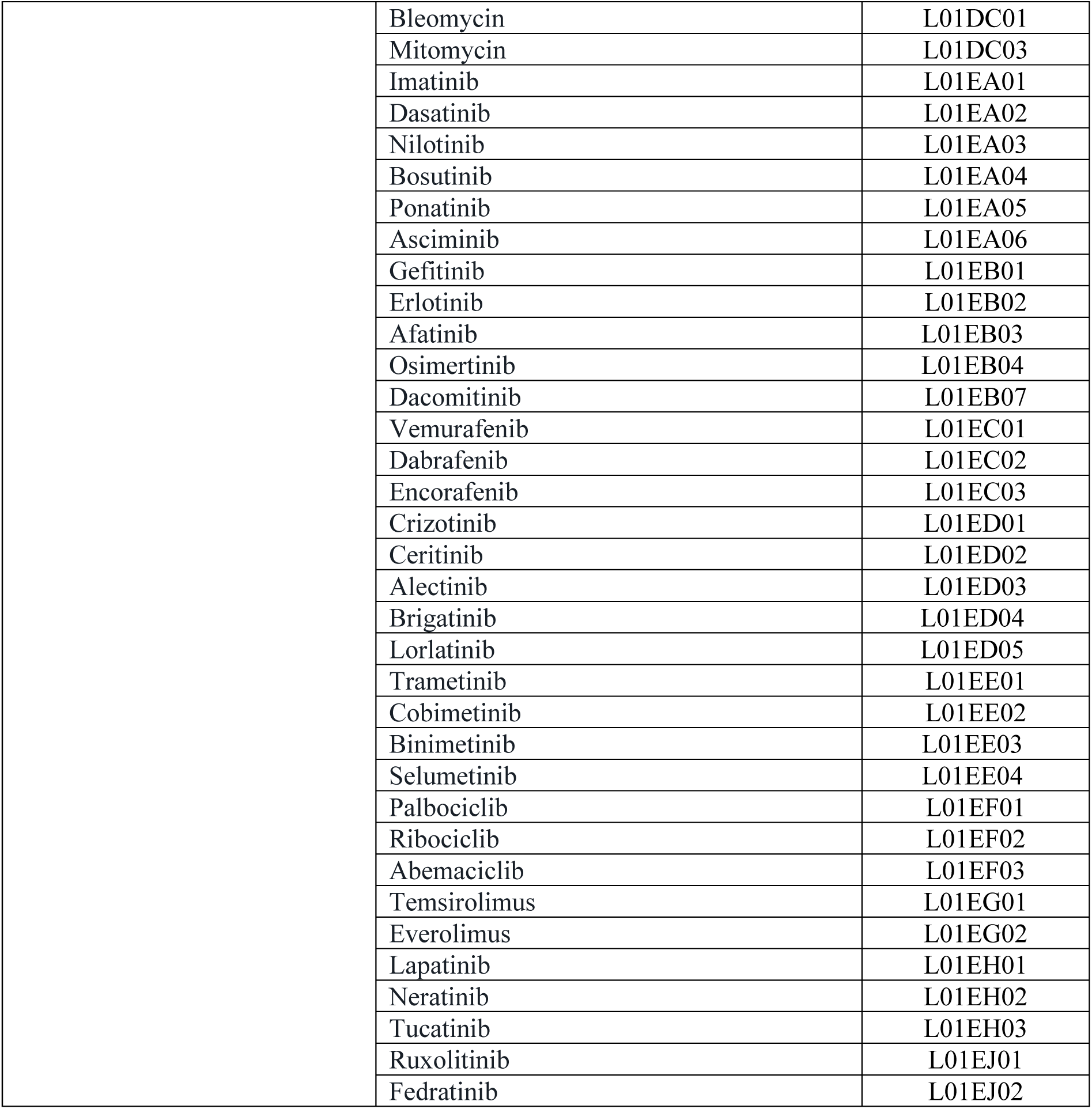
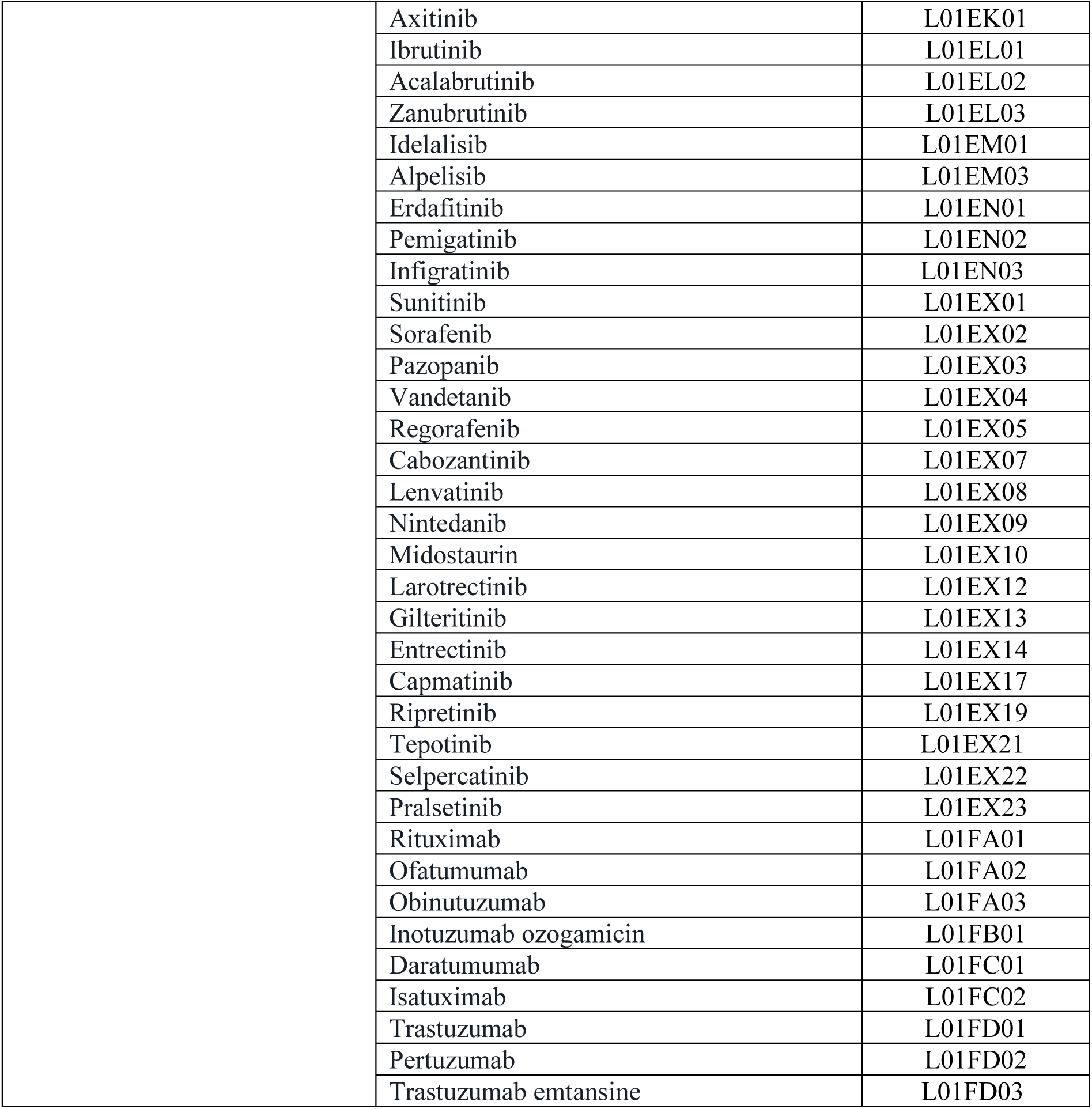
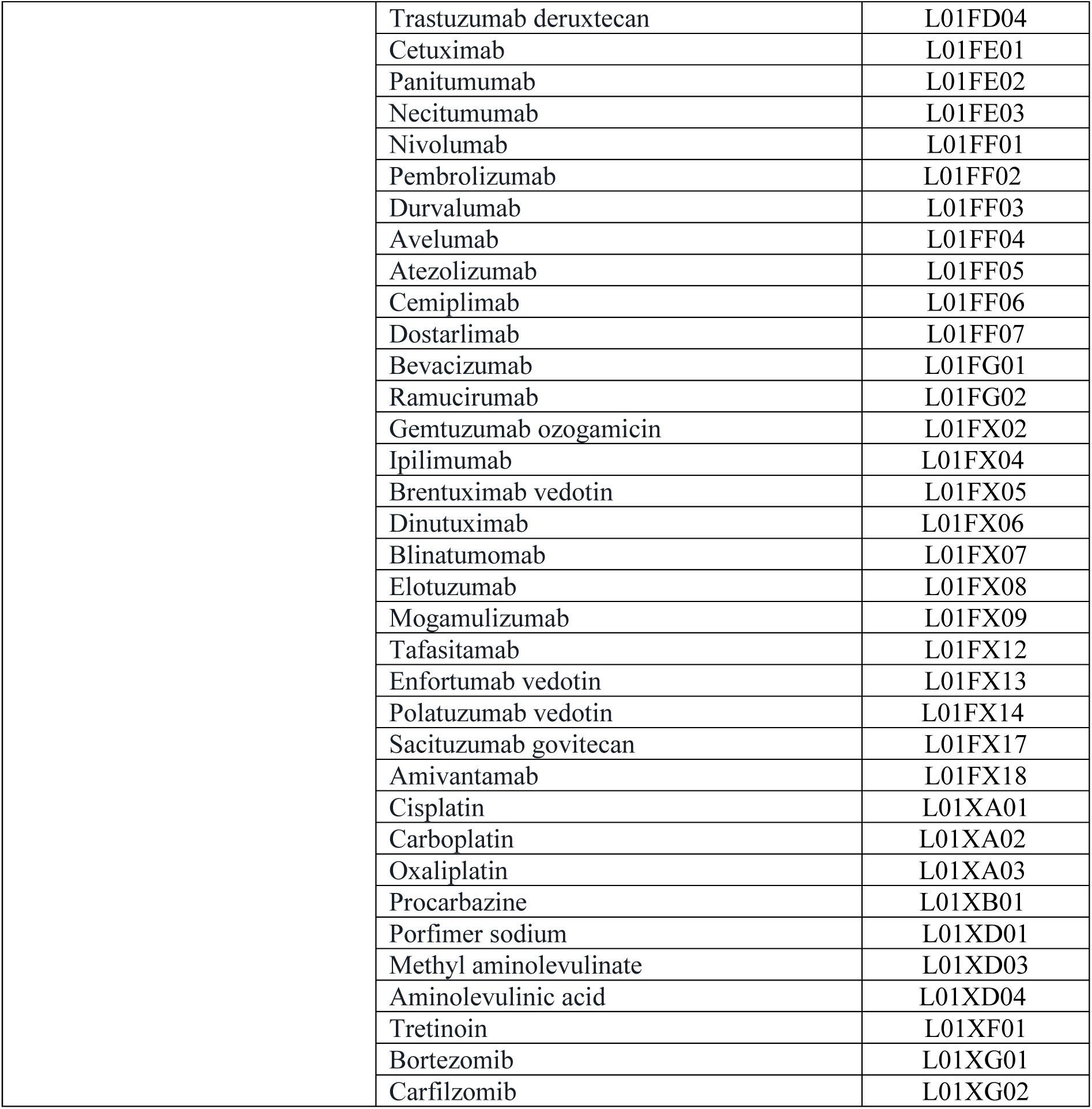
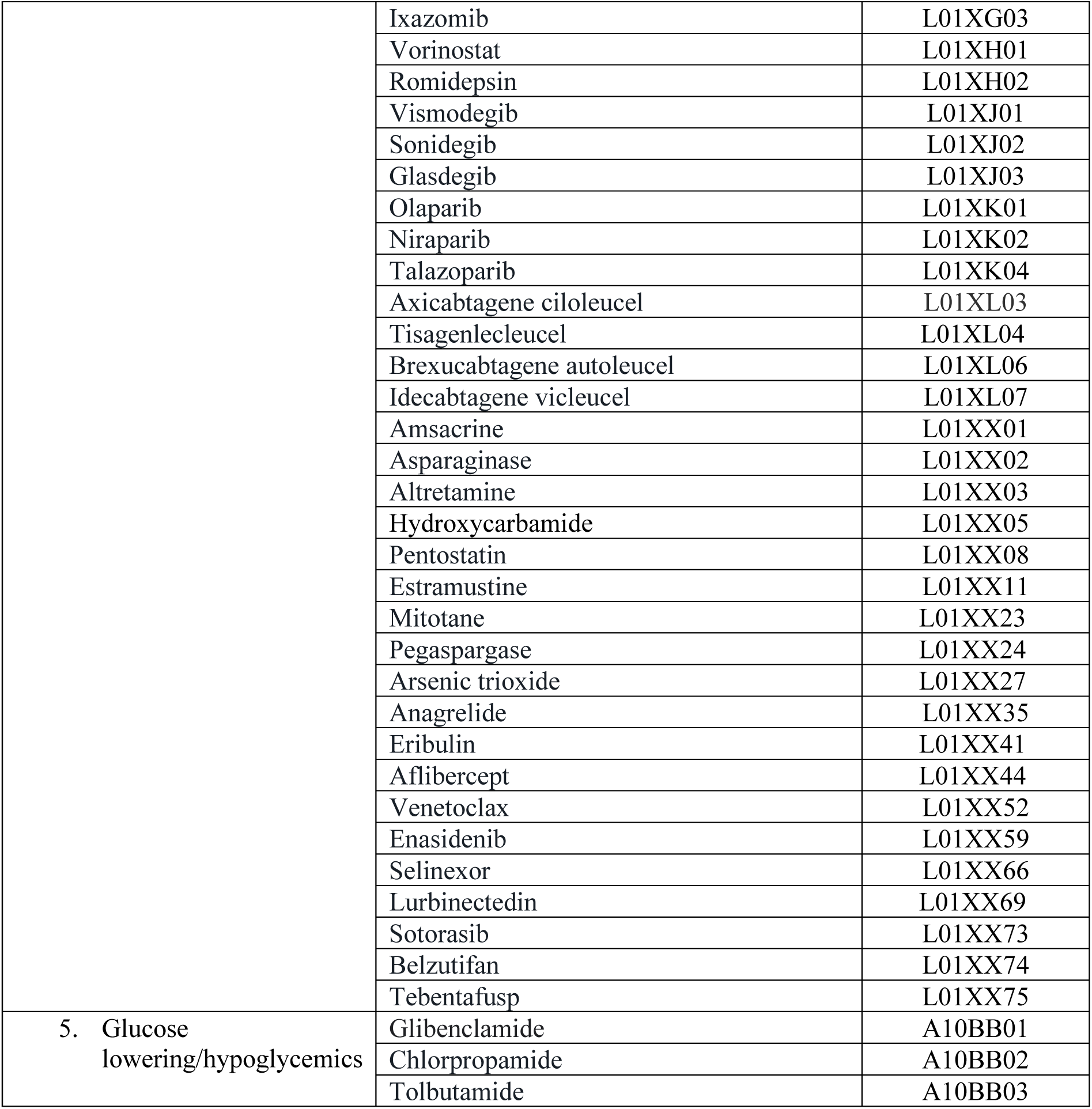
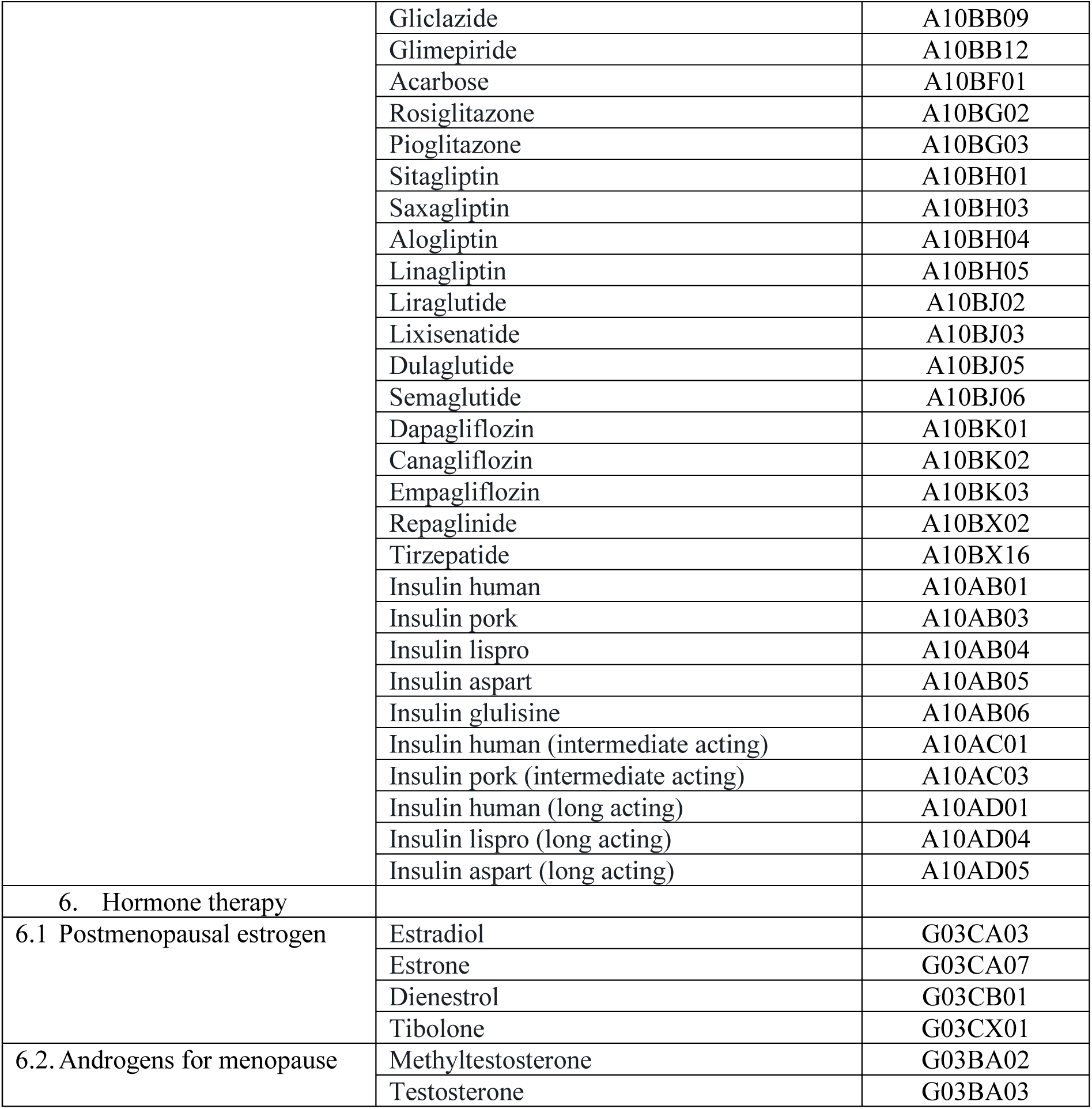
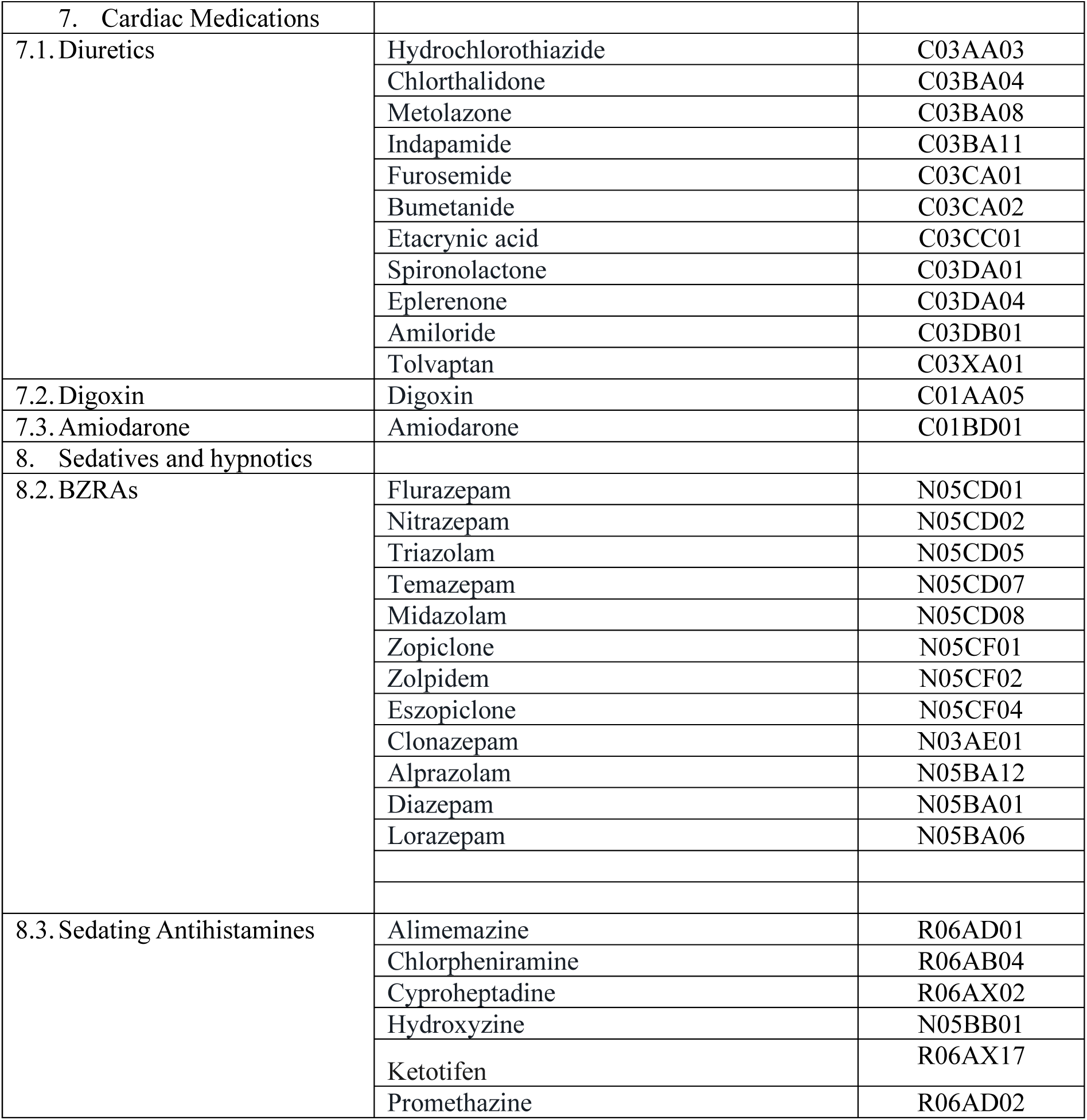
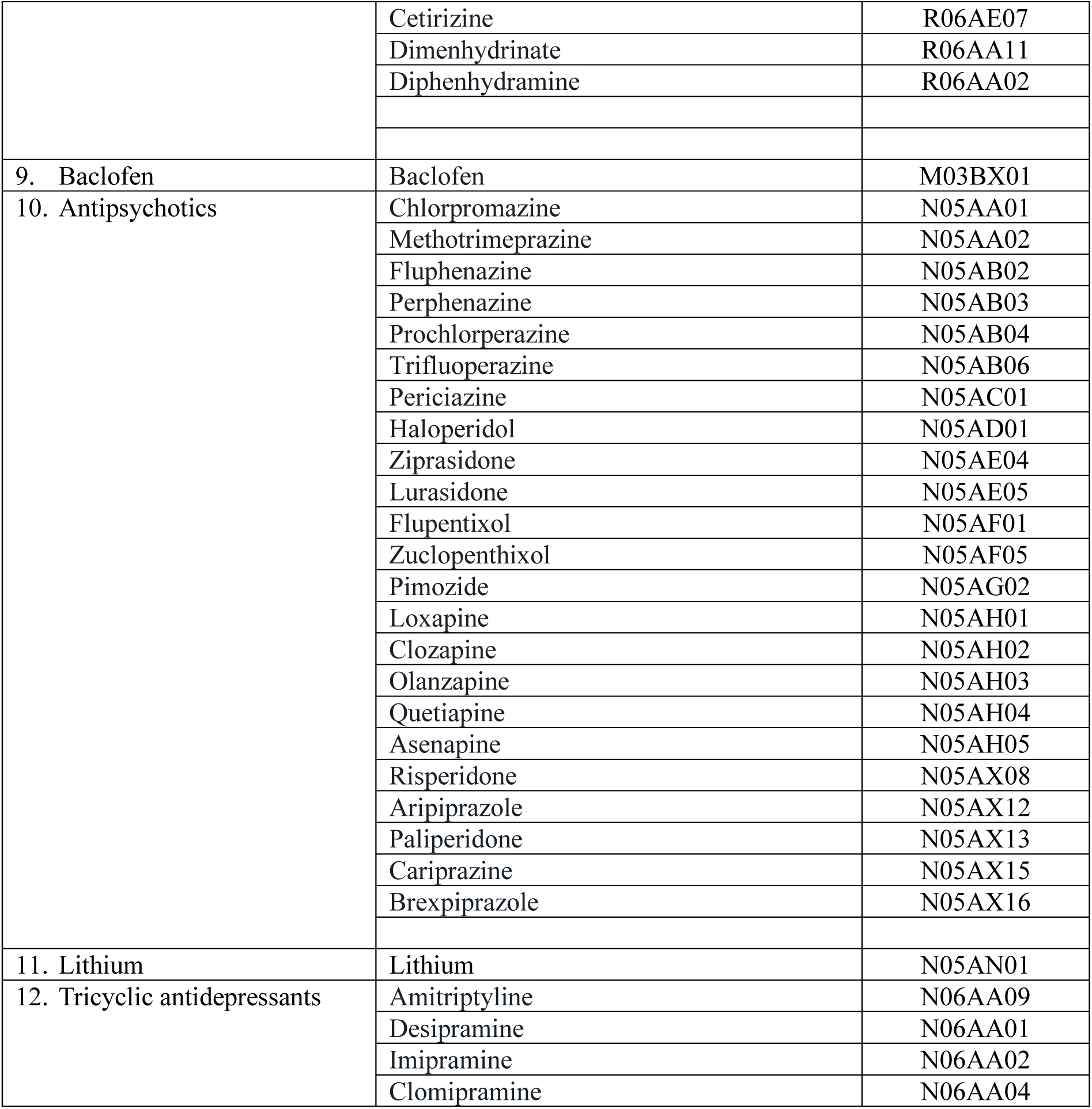
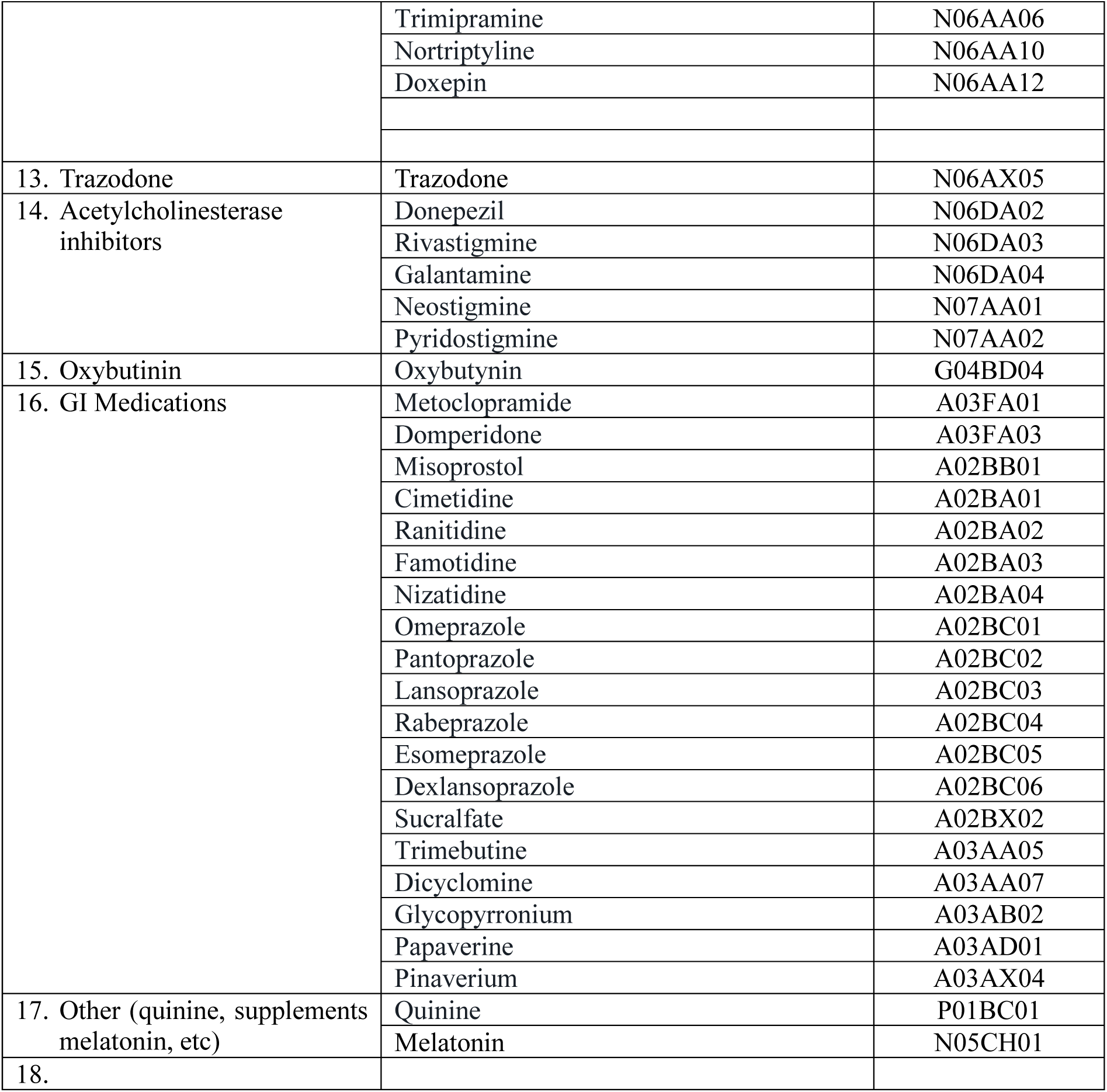
**IMPROVE-IT HRM High-Risk Medications**^27, 28^

**Appendix 2.**
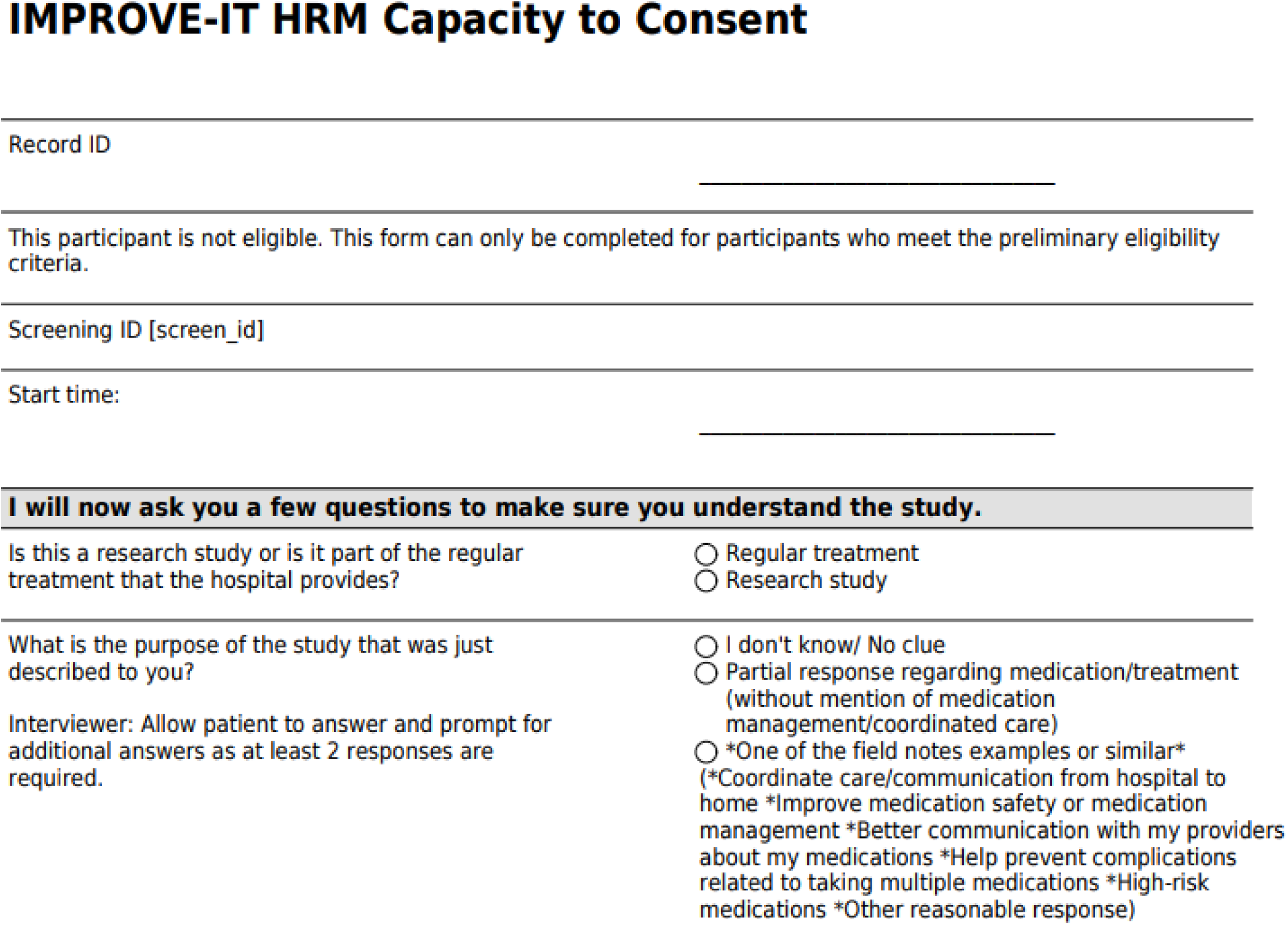
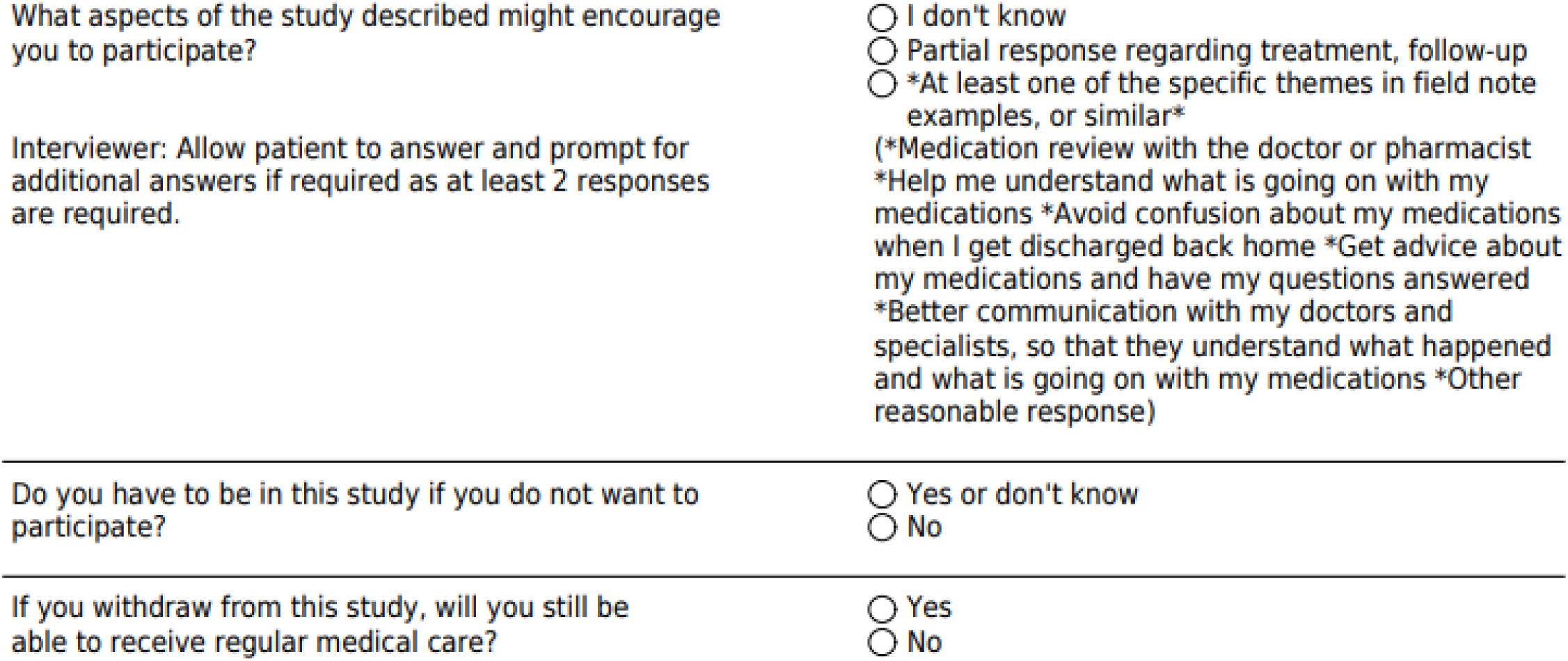
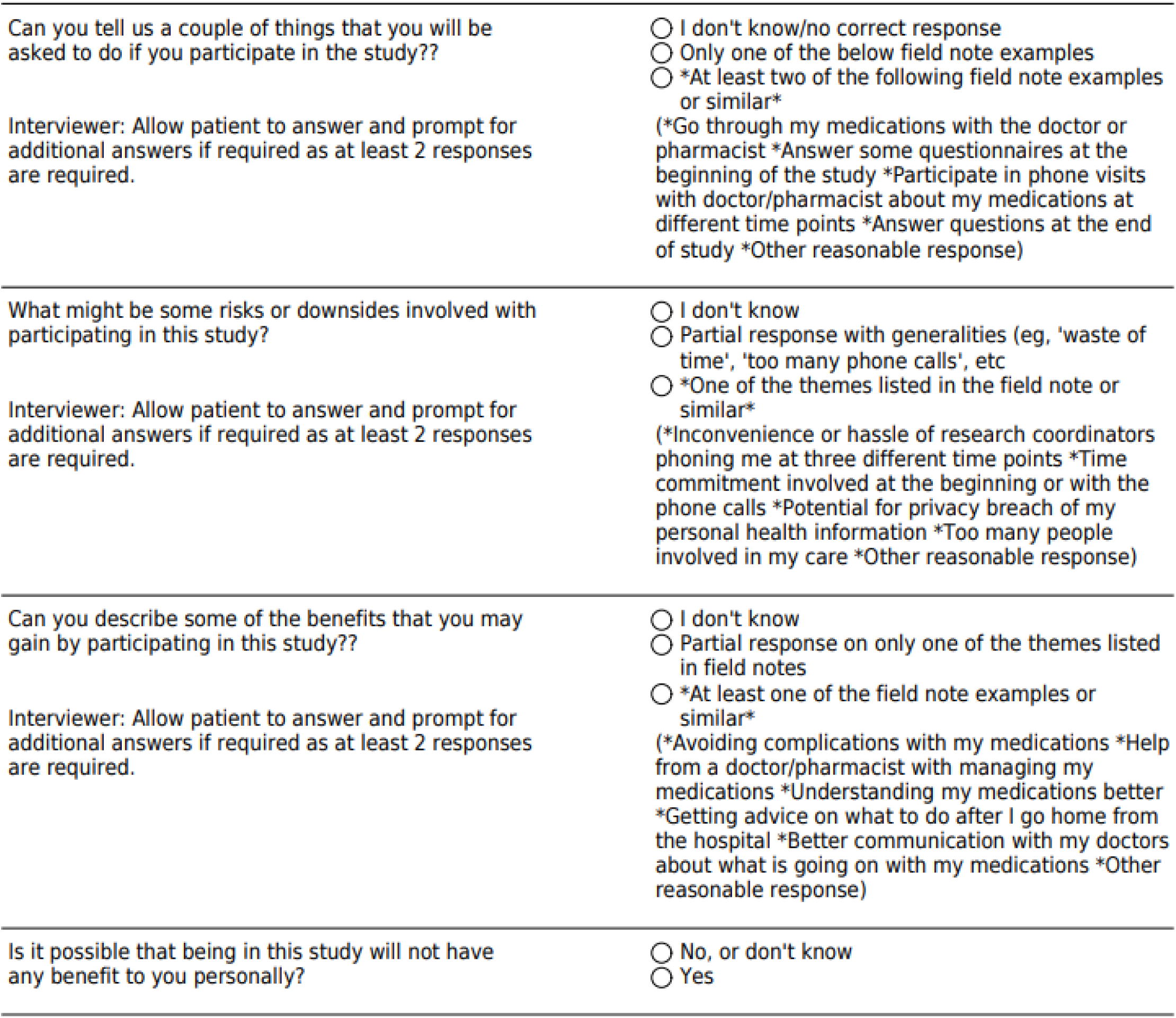
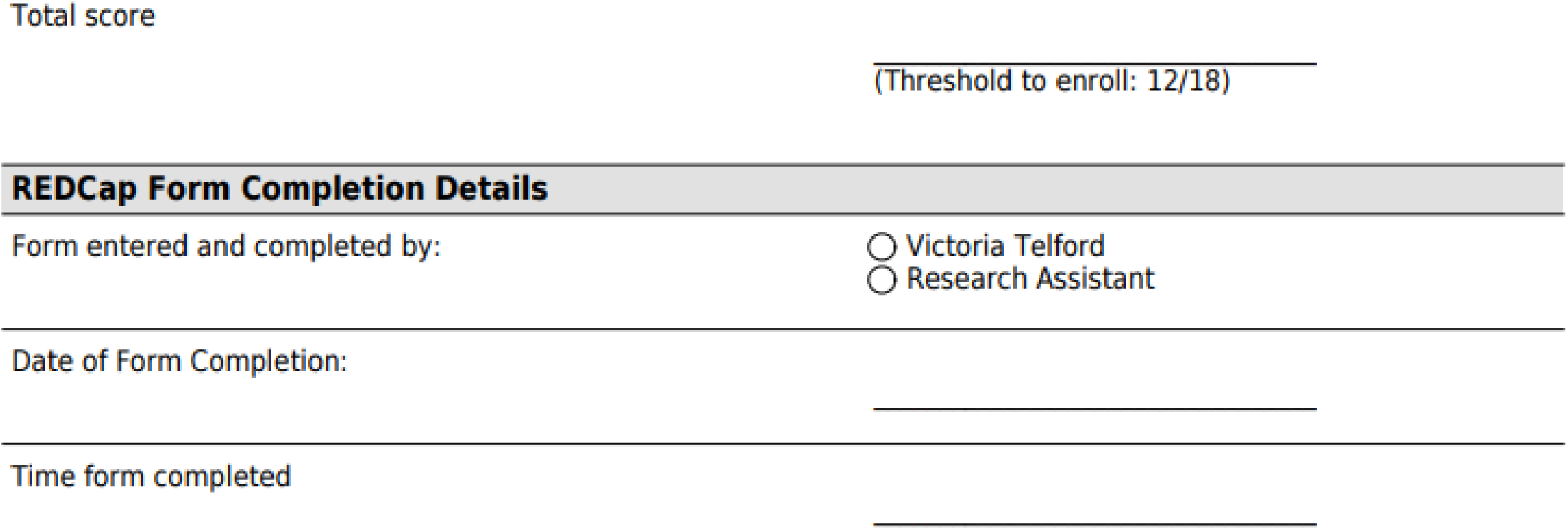
**IMPROVE-IT HRM Capacity to Consent Questionnaire**

## Notes

### Competing Interest Statement

The authors have declared no competing interest.

### Clinical Trial

Clinical Trial ID NCT04077281

### Funding Statement

This study was funded by the Canadian Institutes of Health Research, Funding Reference Number TEG-165595.

### Author Declarations

Hamilton Integrated Research Ethics Board of Hamilton Ontario gave ethical approval for this work (study #7598).

